# Insights into the Mechanisms and Structure of Breakage-Fusion-Bridge Cycles in Cervical Cancer using Long-Read Sequencing

**DOI:** 10.1101/2023.08.21.23294276

**Authors:** Isabel Rodriguez, Nicole M. Rossi, Ayse Keskus, Yi Xie, Tanveer Ahmad, Asher Bryant, Hong Lou, Jesica Godinez Paredes, Rose Milano, Nina Rao, Sonam Tulsyan, Joseph F. Boland, Wen Luo, Jia Liu, Tim O’Hanlon, Jazmyn Bess, Vera Mukhina, Daria Gaykalova, Yuko Yuki, Laksh Malik, Kimberley Billingsley, Cornelis Blauwendraat, Mary Carrington, Meredith Yeager, Lisa Mirabello, Mikhail Kolmogorov, Michael Dean

## Abstract

Cervical cancer is caused by human papillomavirus (HPV) infection, has few approved targeted therapeutics, and is the most common cause of cancer death in low-resource countries. We characterized 19 cervical and four head and neck cell lines using long-read DNA and RNA sequencing and identified the HPV types, HPV integration sites, chromosomal alterations, and cancer driver mutations. Structural variation analysis revealed telomeric deletions associated with DNA inversions resulting from breakage-fusion-bridge (BFB) cycles. BFB is a common mechanism of chromosomal alterations in cancer, and this is one of the first analyses of these events using long-read sequencing. Analysis of the inversion sites revealed staggered ends consistent with exonuclease digestion of the DNA after breakage. Some BFB events are complex, involving inter- or intra-chromosomal insertions or rearrangements. None of the BFB breakpoints had telomere sequences added to resolve the dicentric chromosomes and only one BFB breakpoint showed chromothripsis. Five cell lines have a Chr11q BFB event, with *YAP1/BIRC2/BIRC3* gene amplification. Indeed, *YAP1* amplification is associated with a 10-year earlier age of diagnosis of cervical cancer and is three times more common in African American women. This suggests that cervical cancer patients with *YAP1*/*BIRC2*/*BIRC3*-amplification, especially those of African American ancestry, might benefit from targeted therapy. In summary, we uncovered new insights into the mechanisms and consequences of BFB cycles in cervical cancer using long-read sequencing.

## Introduction

Cervical cancer ranks fourth in worldwide cancer prevalence (Schiffman et al., 2016). Countries with low human development index (HDI) have higher cervical cancer incidence and mortality (Lin et al., 2021) and account for nearly 90% of all cervical cancer deaths annually (Bray et al., 2018; Schiffman and Castle, 2005). Cervical cancer is predominantly caused by oncogenic Human Papillomavirus (HPV) infection, detected in over 90% of cervical cancer patients). (Schiffman et al., 2016). Two main hrHPV types, HPV16 and HPV18 are responsible for at least 70% of cervical cancers (Walboomers et al., 1999).

After infection, the double-stranded circular HPV genome, which is approximately 7900 base pairs (bp) in length, relocates to the nucleus, where it replicates as an extrachromosomal episome. The main drivers of HPV pathogenesis are the E6 and E7 viral oncoproteins. Upon expression, E6 binds to and inactivates tumor suppressor p53, and E7 suppresses the cell cycle regulator pRB (Munger et al., 1992). In many HPV-driven tumors, It is common for HPV to integrate into the human genome, frequently deleting the E1 and E2 genes (Jeon et al., 1995). Since E2 is a transcriptional repressor of E6 and E7, integration results in higher E6 and E7 expression, promoting tumor progression (Munger et al., 2004). HPV integrates into many locations in the human genome, with 37 known hotspots (Warburton et al., 2018). Integration may delete, amplify, or activate cellular genes (Akagi et al., 2014) (Parfenov et al., 2014; Shukla et al., 2014).

Phylogenetic analysis groups each HPV type into lineages and sublineages based on nucleotide variation (Burk et al., 2013). The distribution of lineages and sublineages varies by geographic region and genetic background (Clifford et al., 2019). Cancer risk also varies by lineage and sublineage. For example, the HPV16 A4, D2, and D3 sublineages are more carcinogenic (Mirabello et al., 2016) than the most frequent HPV16 A1 sublineage (Clifford et al., 2019).

Another factor that influences the risk of HPV infection and cancer progression is genetic variation of the host immune system. Thus, the immune response to viral infection is mediated by the Major Histocompatibility Complex (MHC) proteins (Naranbhai and Carrington, 2017). MHC Class I genes, also known as Human Leukocyte Antigens (HLA) A, B, and C, encode glycoproteins found on the surface of all nucleated cells. These complexes bind and present viral peptides to cytotoxic T cells, specialized immune cells that can recognize and kill virally infected cells (Martin and Carrington, 2013). Indeed, most HPV infections are cleared within 1-2 years (Rodriguez et al., 2008). However, some HPV infections can persist and progress to cervical cancer due to various factors interfering with MHC Class I-mediated immune recognition. HLA Class I genes are highly polymorphic, and 90% of people are heterozygous for a given HLA gene (Carrington et al., 1999). This increases the diversity and coverage of the immune response against HPV. However, many cervical cancer cells show loss of heterozygosity (LOH) for the HLA Class I genes (Martinez-Jimenez et al., 2023). This reduces the number and variety of viral peptides that can be presented to CTLs, allowing the virus to evade immune detection. Another factor that impairs the MHC Class I-mediated immune recognition is the expression of HPV proteins that modulate the cellular pathways involved in antigen processing and presentation. For example, the HPV E5 protein inhibits the transport of HLA Class I protein complexes to the cell surface (Miyauchi et al., 2023), and the E6 and E7 proteins prevent apoptosis, which is a process that induces the release of viral antigens and stimulates the immune response (Li et al., 2005). Moreover, the integration of HPV DNA into the host genome disrupts the expression of viral proteins, especially E2, which regulates the transcription of other viral genes (Martinez-Jimenez et al., 2023). This reduces the availability of viral peptides for MHC Class I presentation.

Cervical cancer, like all solid tumors, is characterized by specific chromosomal rearrangements that are vital somatic events during the tumorigenesis process (Berger et al., 2018). The deletion of the telomere can lead to Breakage-Fusion-Bridge (BFB) cycles. This mechanism was first proposed by Barbara McClintock in maize (McClintock, 1941) and involved the deletion of the telomere of a chromosome can lead to Breakage-Fusion-Bridge (BFB) cycles. Telomere loss leads to the formation of a dicentric chromosome that is pulled apart during cell division, resulting in breakage at a random site. BFB cycles can repeat indefinitely, generating complex chromosomal rearrangements and amplifications (Toledo, 2020). In cancer cells, BFB cycles can increase the copy number of segments of the unstable chromosome that contain oncogenes, providing a selective advantage for cells carrying them (Luebeck et al., 2020; Marotta et al., 2017). However, the structure and origin of BFB events in cancer cells are poorly investigated due to the limitations of the conventional short-read sequencing methods (Newell et al., 2022). Long-read whole-genome sequencing (WGS) is a powerful tool for studying BFB and provides an opportunity to understand this mechanism of oncogene activation.

Cervical cancer requires a better understanding of the molecular defects, such as BFB, that drive its progression and resistance. The main treatment options for cervical cancer are surgery, chemotherapy, and radiation. There is currently no targeted therapy approved for cervical cancer. But recent data suggest that 15-20% of patients respond to immunotherapy with checkpoint inhibitors (Chung et al., 2019). Plus, engineered T-cell therapy was recently proven successful in a clinical trial (Nagarsheth et al., 2021). Therefore, the characterizing this cell line panel will provide a valuable resource for the preclinical evaluation of targeted therapy and the further translation of such data into clinical practice.

## Results

### Establishment of a cervical cancer cell line panel

To create a platform to study HPV and cervical cancer, we collected a comprehensive panel of 19 cervical cancer cell lines, and four HPV-positive head and neck squamous cell carcinoma (HNSCC) lines and xenografts. We performed long-read WGS with and without adaptive sampling and full-length cDNA or direct RNA transcriptome sequencing on all cell lines (**Supplemental Figure 1**). **Table 1** summarizes the HPV types, histology, ancestry, age, and HPV integration status of the cell lines. **Figure 1A** shows the diverse ancestry of the subjects: 52% East Asian, 43% European, and 5% African American. In addition, there are eight different HPV types represented, with HPV16 present in 50% (11/22) of our panel.

**Figure 1.**
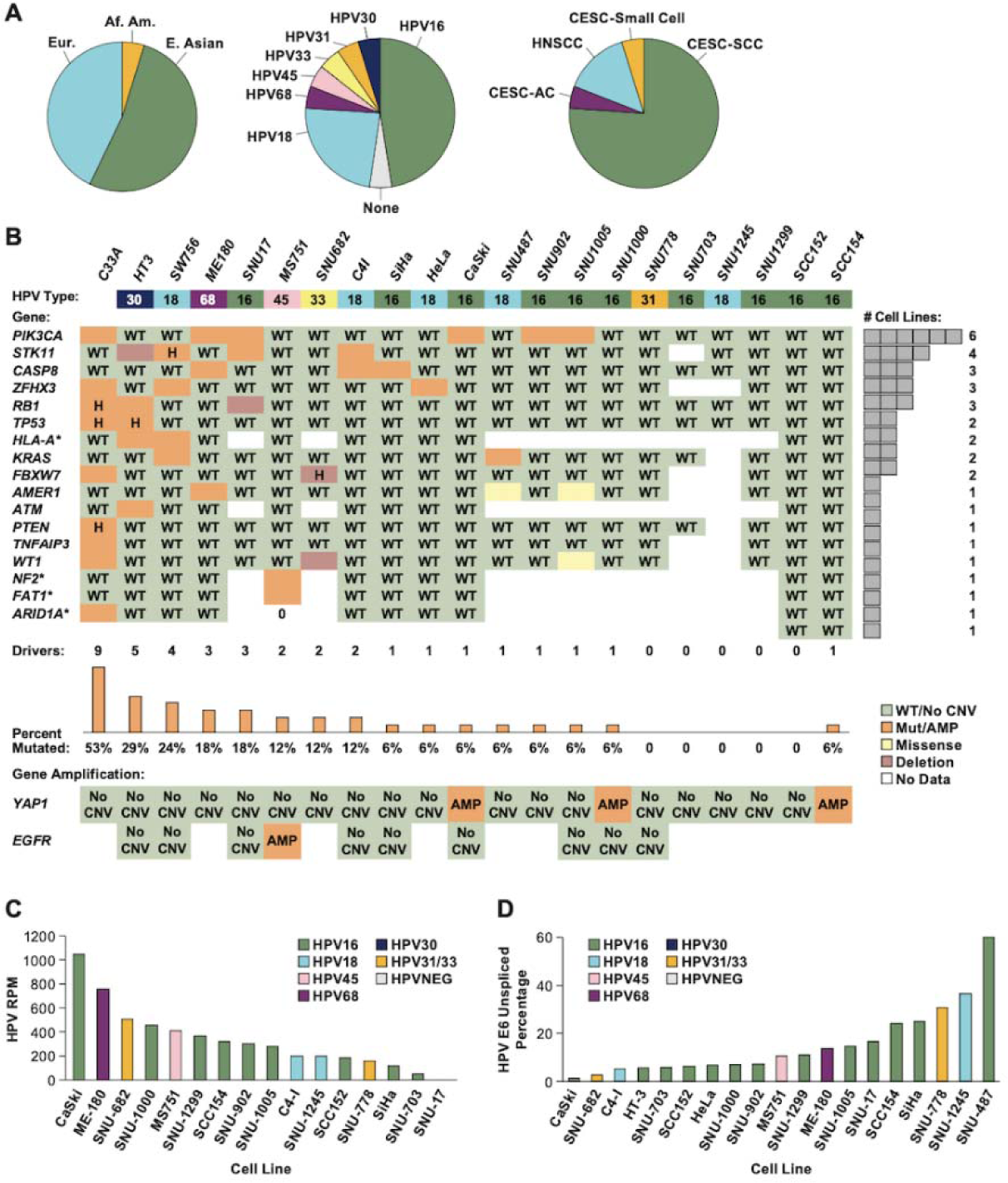
Cell line patient demographics and HPV expression data. **A.** The distribution of the ancestry of the subjects, HPV types, and cancer and histological types among the 22 cell lines is shown. **B.** The cancer driver gene mutations found mutated in at least one cell line are shown. Orange highlights are pathogenic variants and in yellow are variants of unknown impact; H, homozygous variants; WT, wild type; AMP, amplification; no CNV, no copy number variants were found. **C.** The cDNA or RNA reads per million reads mapped (RPM) to the HPV genome is shown. **D.** The percentage of HPV E6/E7 containing reads that are unspliced in the E6 gene.

**Table 1–.**
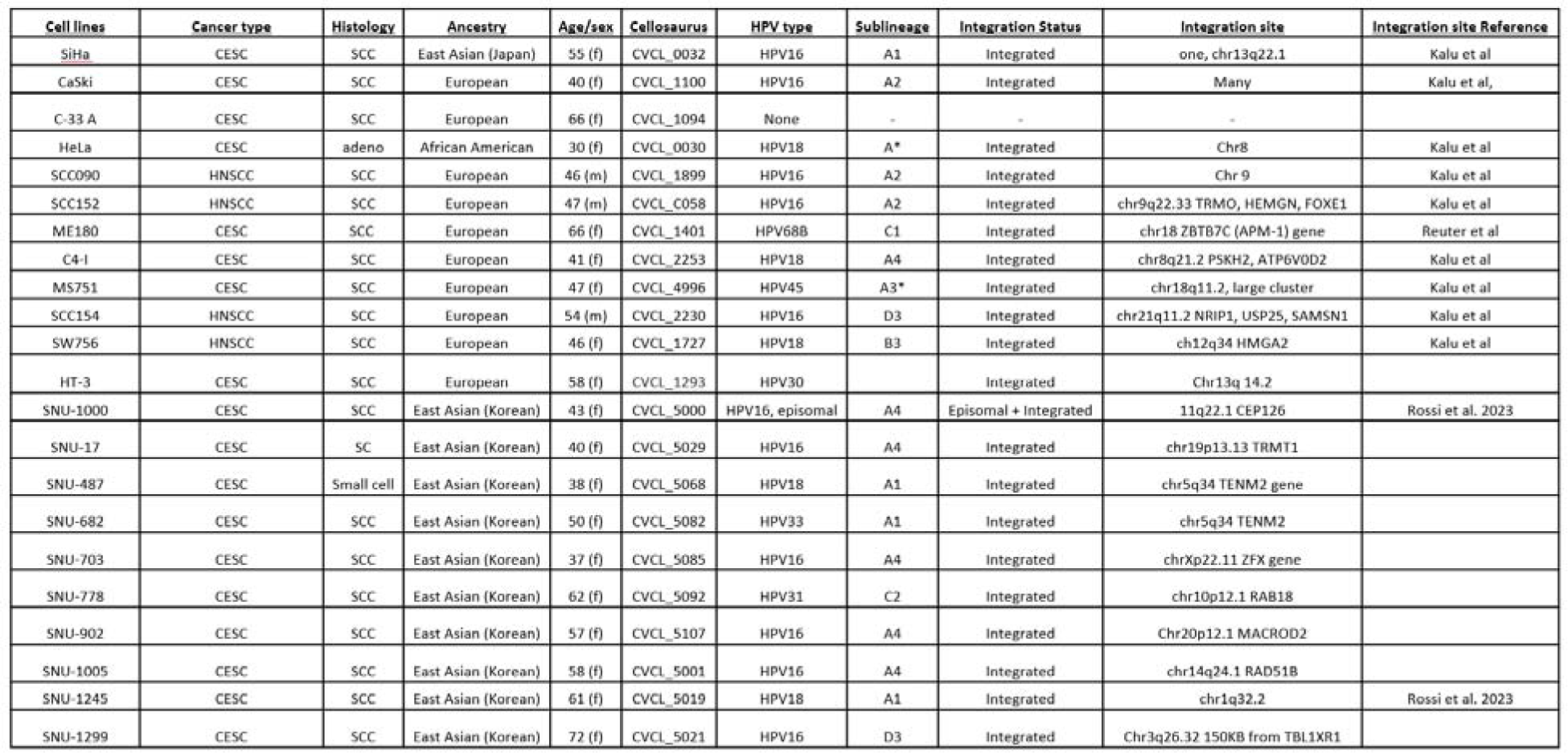
HPV and cervical cancer cell lines and their identifying features. HPV cell line information including HPV type, HPV sublineage, patient demographics, and integration sites are listed. Integration sites were originally identified in the references listed, or in this paper. Characteristics of each cell line were determined utilizing techniques outlined in the methods. For some cell lines (*), the sublineage could not be determined because the sequence length was insufficient. CESC – cervical squamous cell carcinoma and endocervical adenocarcinoma, HNSCC – head and neck squamous cell carcinoma.

We performed whole genome sequencing using ligation sequencing on the Oxford Nanopore platform. We used sheared DNA, which generated 8 to 14 million reads with an N50 value of 9 to 11 kb, and unsheared DNA, which generated 8 million reads with an N50 value 30 kb). Each DNA sample was sequenced on a single MinION flow cell (**Supplemental Tables 1**). We used adaptive sampling to enrich for sequences relevant to HPV and cervical cancer (Kovaka et al., 2021; Loose et al., 2016). These regions included 299 genes that are frequently mutated or altered in cancer, genomic loci that are prone to HPV integration, and 13 hrHPV types. This approach yielded 7-24 million reads on single MinION flow cells, with 5-10x coverage of most sampled genes and HPV integration breakpoints (**Supplemental Table 2**). Rejected reads from adaptive sampling had a mappable length of still have 400-500 bp and were used to generate high-resolution copy number profiles. As PromethION flow cells became available we repeated some cell lines achieving 20-30-fold coverage with an N50 of 10-11 kb (**Supplemental Table 1, 2**).

To identify somatic driver mutations, we analyzed all variants in the 299 frequently mutated genes identified in the TCGA pan-cancer study (Bailey et al., 2018). Previously annotated somatic mutation data for 12 cell lines we studied were available from COSMIC (https://cancer.sanger.ac.uk/cell_lines). We also performed manual variant calling of mutation hotspot and selected genes on previously uncharacterized cell lines (**Figure 1B**). Consistent with prior research on cervical tumors (Cancer Genome Atlas Research et al., 2017; Lou et al., 2015), *PIK3CA* is the most frequently mutated gene in the cell lines, and there are mutations in *STK11*, *CASP8*, *ZFHX3*, *RB1*, *HLA-A*, and *KRAS* in more than one cell line. In total, 16/22 cell lines have mutations in three or fewer driver genes. However, C33A, an HPV-negative cell line (Scheffner et al., 1991), has nine known driver mutations, and HT-3, a cell line with an unknown-risk HPV (HPV30) (Naeger et al., 1999), has five mutations. Therefore, our cell line panel recapitulates the mutational spectrum of primary cervical tumors. Furthermore, this result suggests that cell lines without a hrHPV type require additional driver mutations for cancer progression.

Both direct cDNA and direct RNA methods were used to characterize HPV and cellular gene expression. Direct cDNA sequencing yielded 3-10 million reads on a single MinION flow cell and Direct RNA 0.8-1.2 million reads (**Supplemental Table 1**). The cell lines display a wide range in the level of HPV RNA expression, as determined by reads aligning to the HPV genome per million reads (RPM). The CaSki and ME180 cell lines have the highest HPV expression while SNU17 and SNU703 have the lowest (**Figure 1C**). One of the oncogenic functions of hrHPV E6 protein is to degrade the tumor suppressor p53. However, a specific splicing event within the E6 gene occurs in most tumors and cell lines with hrHPV and deleting a portion of E6 and rendering E6 unable to degrade p53. Furthermore, the E7 protein has been shown to be exclusively translated from this spliced mRNA (Tang et al., 2006)). SNU-487 and SNU-1245 showed the highest frequency of E6 gene splicing at 60% and 36%, respectively, while SNU-682 and CaSki exhibited only 3% and 2% splicing frequency (**Figure 1D**).

### An MHC haplotype is frequently deleted in cervical cancer cell lines

The expression of human leukocyte antigen (HLA) molecules is essential for the recognition and elimination of tumor cells by the immune system. Therefore, we characterized the HLA genotype of our cell lines (**Figure 2A**). A total of 9 out 18 cell lines have a homozygous genotype across all class I and class II HLA genes, indicating a deletion of one MHC haplotype. The percentage of homozygosity for each HLA class I gene was 56% for *HLA-A* and *HLA-B* and 61% for *HLA-C* Figure 2B. This frequency is significantly higher than seen for 159 non-cervical cell lines (X2= 8.0, P=0.0047). (Boegel et al., 2014). In addition, at least two cell lines, SW756 and HT-3, have loss-of-function mutations in the *HLA-A* gene. These results indicate a strong selective pressure to eliminate an MHC haplotype or allele in cervical tumors.

**Figure 2.**
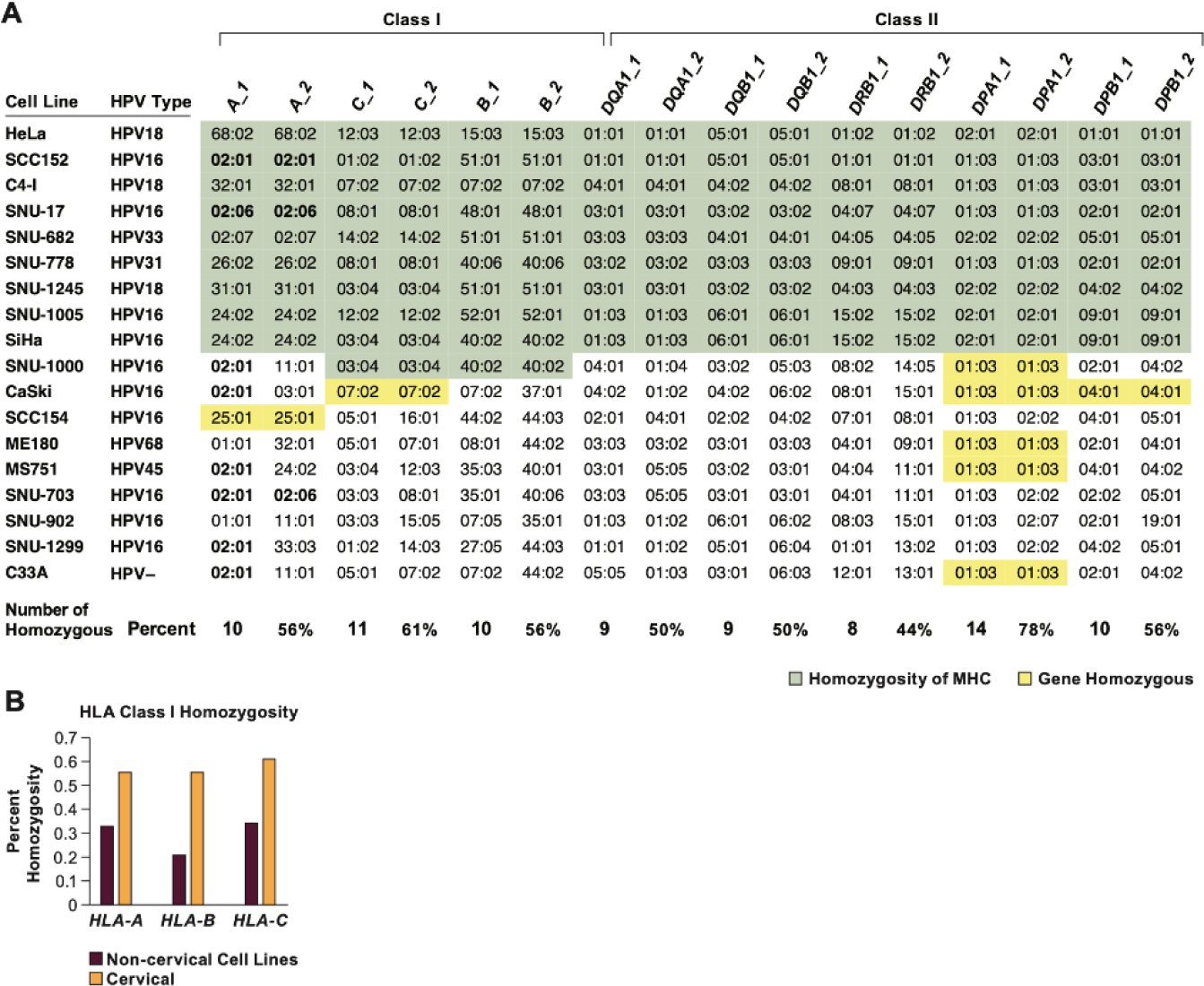
HLA class I homozygosity in cervical cancer cell lines. **A.** The genotype for each class I and II HLA genes is shown with loci homozygous across all or most genes indicated in green, and across 1-3 genes in yellow. The number and percent homozygosity for each gene is indicated at the bottom. **B.** The percent of tumor samples homozygous for individual HLA class I genes. The data for non-cervical cancer cell lines are from (Boegel et al., 2014).

### Location and impact of HPV integration

To determine the spectrum of HPV integration sites, we mapped their locations in the human genome (**Figure 3A, Supplemental Figure 2, Table 1**). As observed in cervical tumors, the HPV integration sites in the cell lines are widely distributed and found on most human chromosomes (Bodelon et al., 2016). Each cell line has a unique integration site, and 10 out of 20 are in HPV integration hotspots. In addition, six cell lines have an integration near a known super-enhancer active in cervical cells (Warburton et al., 2021). In SNU-487 and SNU-682, integration occurs at chromosome 5q34, near the *TENM2* gene. TENM2 mediates cell adhesion and may represent a new cervical cancer hotspot (Bagutti et al., 2003).

**Figure 3:**
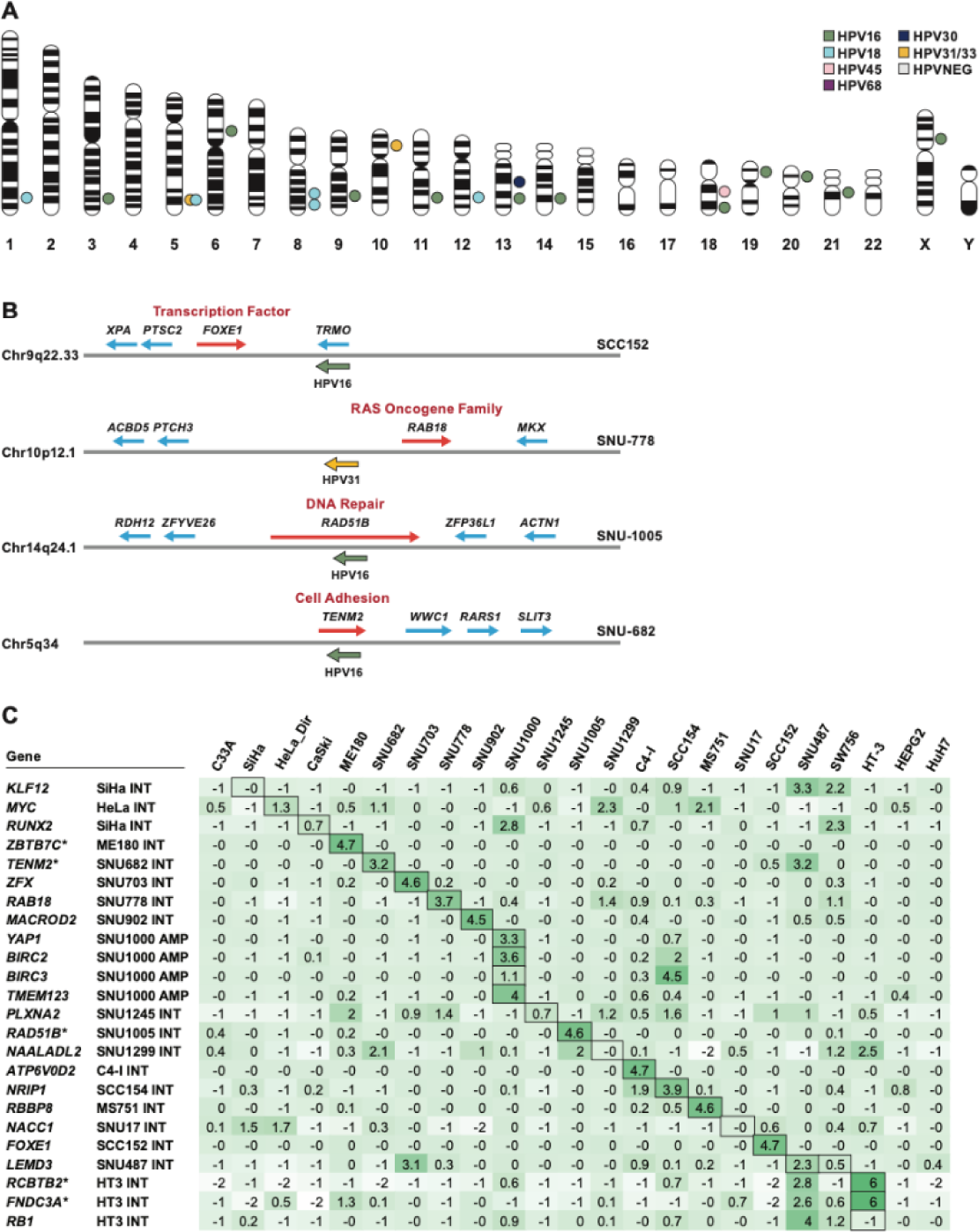
HPV integration and gene expression. **A.** A map of all integration events in the human genome is shown for 20 HPV+ cell lines. Each point represents an integration event, of which there may be one or multiple integration breakpoints. For the cell lines CaSki and SCC152, with multiple integrations, only the transcriptionally active site is shown. Each point lies in an approximate location of the integration event. The exact locations are included in Table 1 and **Supplementary Table 2. B**. Diagram of the HPV integration locus and flanking genes in four cell lines. The red arrows indicate a gene at the integration locus that is overexpressed in that cell line. **C.** The relative gene expression of each gene at or near an integration site was calculated as a Z-score; bold boxes are genes that are near the HPV integration. Darker green colors indicate higher values, showing those genes are highly expressed in those cell lines.

To investigate the impact of HPV integration on host genes, we examined the expression level of genes within approximately 1 Mb of integration events and compared them to the same genes in the rest of the cell line panel. We observed that HPV integration activates the expression of one or more genes in the region in almost every cell line. For example, in SCC152, the *FOXE1* transcription factor gene is highly expressed, and in SNU-778, the *RAB18* gene (part of the RAS oncogene family) is overexpressed. The HPV integration occurs within a gene body in several cell lines, and HPV splices into one or more exons. We predict that this will result in insertional inactivation of that gene. For example, in SNU-1005, the *RAD51B* DNA repair gene is inactivated by insertion of HPV31 (**Figure 3B**). We quantified the relative expression of genes near integration sites in all cell lines using a Z-score statistic (**Figure 3C**). Almost every cell line has at least one gene highly expressed relative to the other cell lines without integration at that locus. Many cell lines have integration activating at least one gene of oncogenic importance, such as the *MYC* or *YAP1* oncogenes and the *RUNX2* transcription factor gene.

### Gene expression in cervical cancer cell lines

We also identified oncogenes overexpressed independent of HPV integration. The MS751 cell line has gene amplification and mRNA over-expression of the EGF receptor gene (*EGFR*). Similarly, the *YAP1* oncogene is overexpressed and amplified in SNU1000 and SCC154 cells. *YAP1* encodes a downstream nuclear effector of the Hippo signaling pathway involved in development, growth, repair, and homeostasis (Hatterschide et al., 2022; Pearson et al., 2021) (**Supplementary Figure 3**).

To understand the molecular basis for cervical cancer in the absence of HPV or the presence of unknown-risk HPV types, we analyzed C33A, an HPV-negative cell line, and HT-3, a cell line with the unknown-risk HPV30 virus. C33A cells are homozygous for the pathogenic R273C mutation in *TP53* and homozygous for the c.1961-1 G>A splice site mutation in *RB1*. Therefore, somatic mutations can substitute for the primary oncogenic function of HPV (**Figure 4A, B**).

**Figure 4.**
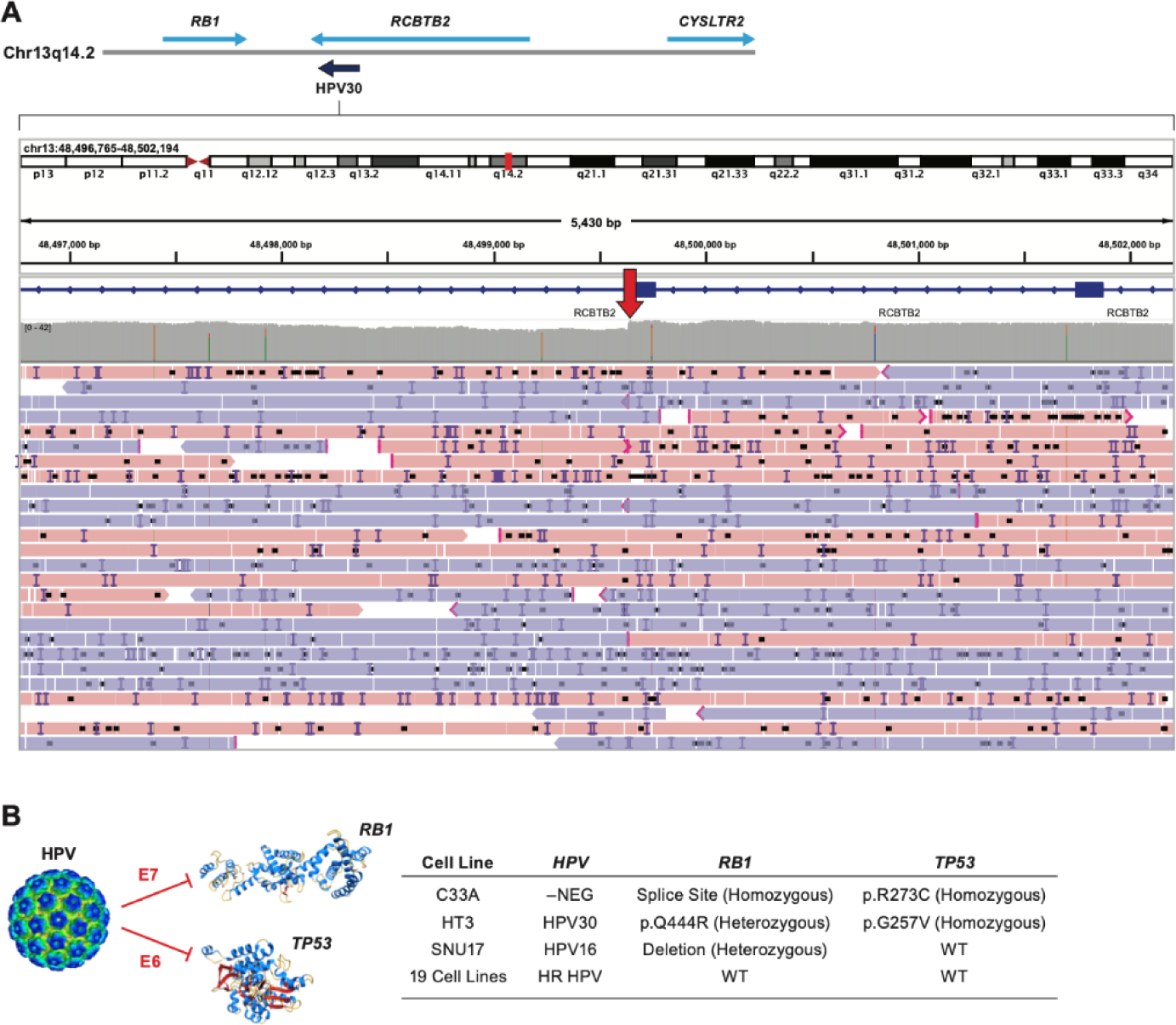
*TP53* and *RB1* mutations in HPV-negative and moderate-risk HPV cell lines. **A.** The Integration site of HPV30 in HT-3 cells demonstrates that the integration is 19 kb 3’ to the *RB1* gene within the *RCBTB2* gene. **B.** A model showing that the HPV E6 and E7 proteins inhibit p53 and pRB respectively and mutations in *RB1* and *TP53* occur almost exclusively in cell lines without HPV or HPV of unknown risk.

In HT-3 cells, long-read WGS revealed that the unknown-risk HPV type, HPV30, is integrated on chromosome 13, 517 kb from the *RB1* gene, within the *RCBTB2* gene. This cell line is also homozygous for a *TP53* mutation (G245V) and is heterozygous for the *RB1* mutation Q444R, known to affect the splicing of exon 13 (Scheffner et al., 1991). In the 20 cell lines with high-risk HPV types, only SNU-17 cells contain a deletion in 1 allele of *RB1*, and none have a *TP53* mutation (**Figure 1B**, **4B**). Therefore, somatic mutations in the *TP53* and *RB1* genes can result in cervical cancer with unknown/low-risk HPV types or the absence of HPV.

### Breakage-Fusion-Bridge events identified in cell lines

We generated copy number plots for each chromosome using the Oxford Nanopore (ONT) DNA reads for each cell line. We observed that some chromosomes showed a focal gain in coverage, followed by a steep drop in coverage and loss of heterozygosity near the telomere (**Supplemental Figure 4**). Junction analysis identified inverted sequences consistent with BFB events (**Figure 5**). Additionally, we used we used Severus (Methods; manuscript in preparation) to identify inversions characteristic of BFB. We found 13 BFB events in nine cell lines (**Supplemental Table 3**).

**Figure 5.**
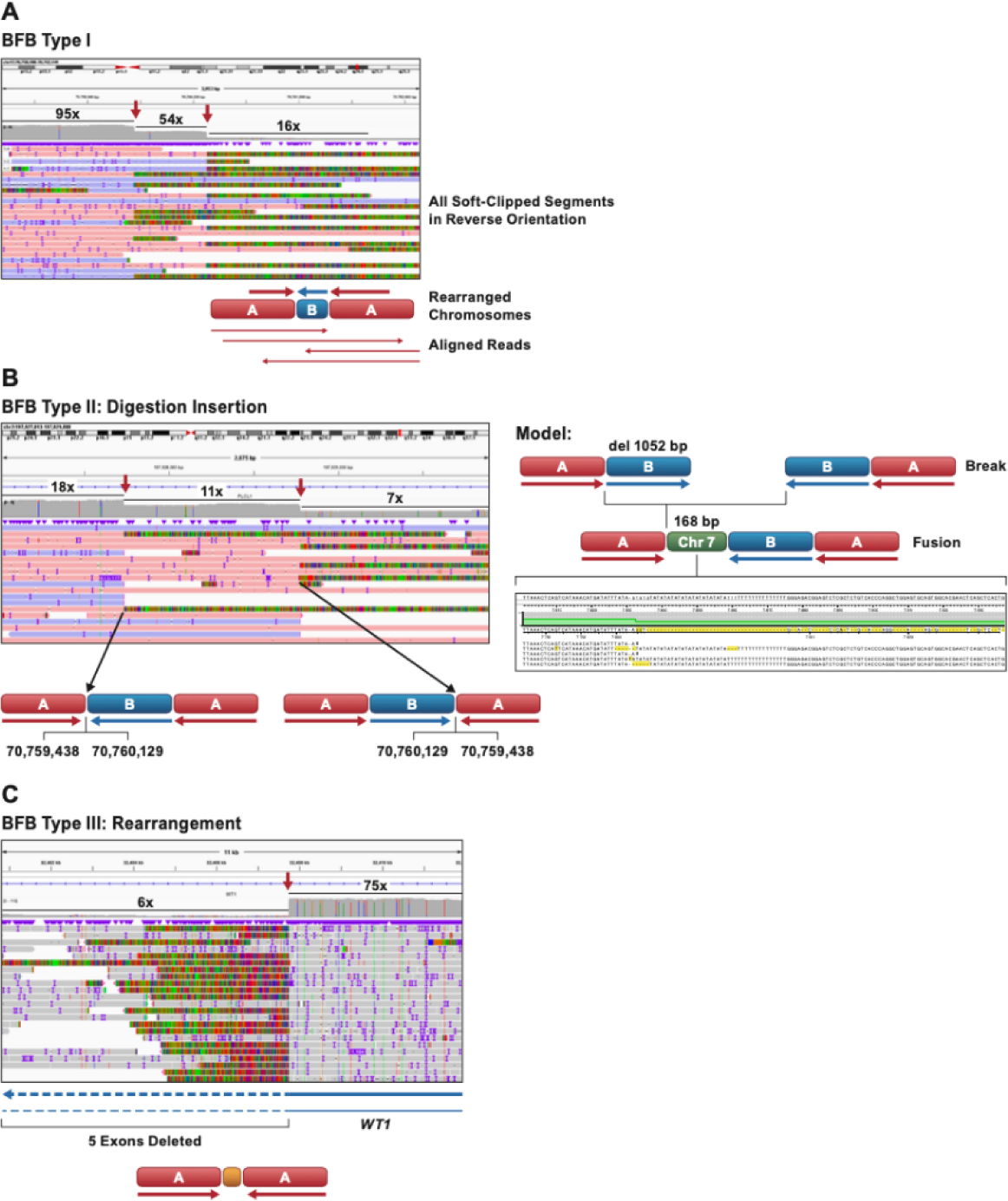
Structure of BFB events. **A.** The alignment of reads at the inversion site of a Type I BFB event on Chr17q in HT-3 cells is shown. The coverage drops from 95x (segment A) to 54x (segment B) to 16-fold (reads from non-rearranged allele). All soft-clipped portions of reads are inverted in relation to the aligned portion of the reads. Arrows mark the start and end of the segment shown as B in the diagram below. All the reads spanning the junction are consistent with the fusion of inverted chromatids with staggered ends. **B.** A proposed model for the formation of the BFB junction of chromosome 2 in SNU-1000 cells, a Type II BFB event. Following the deletion of one of the ends (del 1052 bp), a segment of chromosome 7 is inserted in between the joined chromosomes. The coverage drops from 52x (segment A) to 33x (segment B) to 11x. **C.** A Type III BFB event on chromosome 11p in SNU-682 cells, inside the *WT1* gene. The coverage drops from 75x to 6x. The junction contains a complex sequence likely derived from sequences adjacent to the breakpoint (see **Supplemental Figure 4G**).

To explore the molecular mechanism underlying the formation of BFB events we analyzed the long-read DNA sequences at the junction of the amplification events. At each junction site there is a stepwise drop in coverage along with inverted reads. **Figure 5A** shows an example of a telomere deletion at chromosome 17q24.3 in HT-3 cells. We found that the junctions were composed of inverted reads with different lengths, indicating that one of the chromatids had a deletion before joining with the other. This is consistent with the model shown in **Figure 5A** where the copy number of the B segment is half that of segment A. We propose that segment B was generated by a breakage-fusion-bridge (BFB) cycle involving a breakage and duplication of one chromatid during anaphase, followed by nuclease digestion of the free ends and fusion of the resulting staggered ends. In the HT-3 Chr17q BFB event, the size of the B segment was 688 base pairs. We observed a total of 13 BFB events in 9 of the cell lines, which we termed and classified those into three types based on the origin and structure of the DNA segments at the junctions.

Type I BFB events (**Figure 6A**) were similar to the one described above, with junction segments from the same chromosome. We identified eight Type I BFB events in six cell lines (**Supplemental Table 4**). Type II BFB events (**Figure 5B**) involved insertion of a segment from different chromosomes inserted during fusion process. We found two Type II BFB event in two cell lines. Type III BFB events (**Figure 5C**) involved segments from the same chromosome that were rearranged before or during the fusion process. We detected two Type III BFB events in two cell lines. One of the Type III events affected the *WT1* tumor suppressor gene in SNU-682 cells, causing a deletion of 5 exons and a loss of *WT1* expression (**Figure 5C**). SNU-1000 cells have a complex rearrangement on Chr11 involving HPV sequences (Rossi et al., 2023). To our knowledge, this is the first description of a BFB event leading to the inactivation of a tumor suppressor gene in cancer. We also attempted to characterize the structure of the DNA centromeric to the BFB junctions. In some cases, we observed additional inversions and rearrangements, such as in SNU-703 Chr11 (**Supplemental Figure 5C**), while in other cases, we noticed a gradual decrease in read coverage.

**Figure 6.**
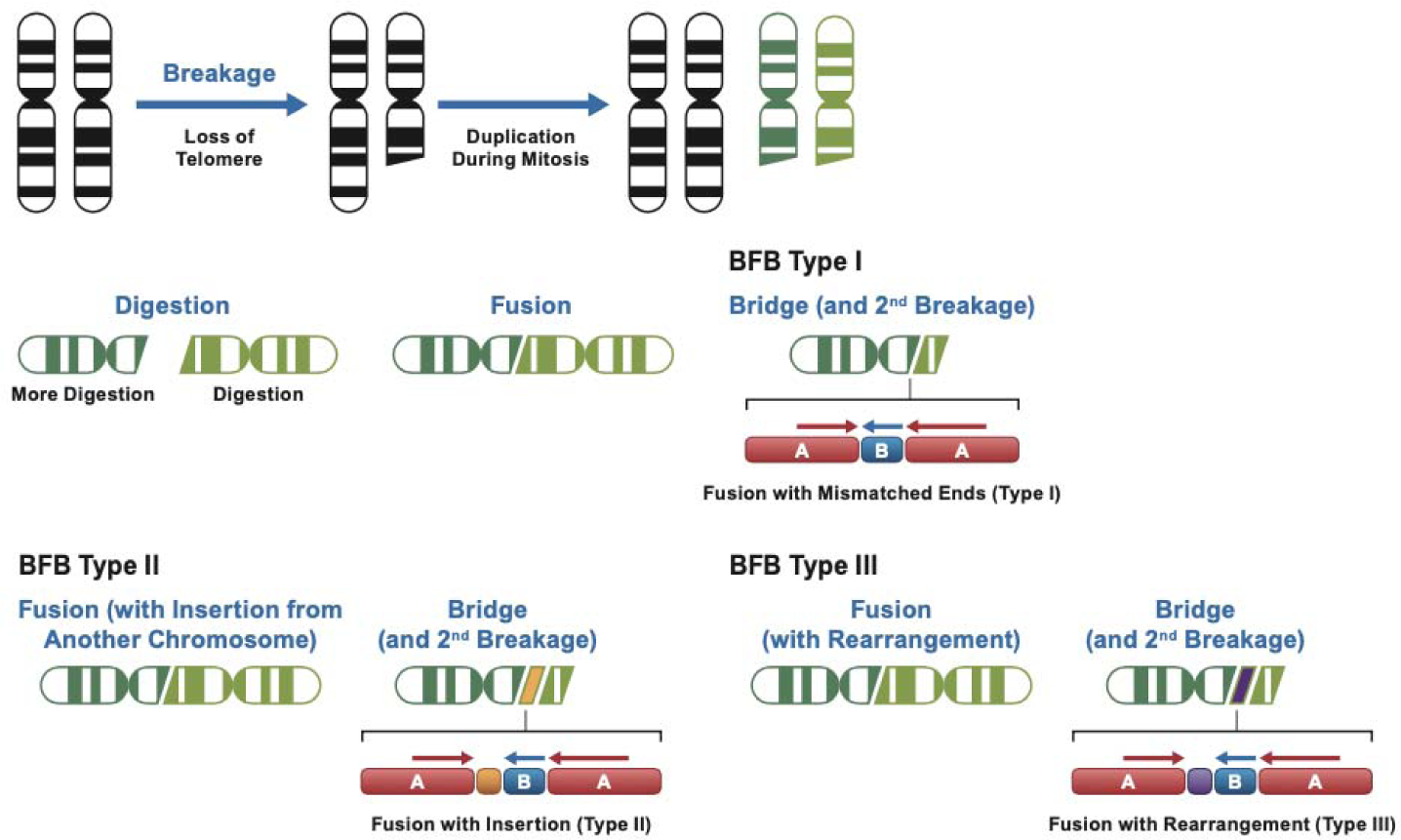
Model for BFB types. Model for a BFB Type I event. A telomere deletion results in a pair of deleted chromatids during mitosis. The free ends are subject to exonuclease digestion, and uneven digestion generates staggered ends. Fusion results in a lower copy number (one half) of the B segment. Type II events are formed when a segment from another chromosome (orange) is inserted at the junction, and Type III events involve insertion of a sequence derived from the sequences flanking the breakage site (purple).

### Canonical telomeres are not found at Breakage-Fusion-Bridge sites

The internal architecture of the BFB amplifications is defined by breakpoints with foldback inversions and sequence digestions. However, it is still unclear how BFB amplifications affect the overall karyotype. We hypothesized that if the derived chromosome would need to acquire a telomere to reach a stable state after one or several rounds of BFB amplifications (Kwon et al., 2020). To explore this hypothesis, we generated haplotype-specific coverage plots of the four cell lines with the highest sequencing coverage (CaSki, SNU-1000, SCC152, and HT-3). In all five BFB events deleted in these cell lines we observed reduced coverage on the telomeric side of BFB, compared to the centromeric side. In CaSki the telomeric side was also characterized by loss of heterozygosity (**Figure 7D**). This suggests that the chromosomal sequence directly after the BFB event on the telomeric side was lost in all cases. In 3 cases that did not have loss of heterozygosity, but instead had reduced telomeric coverage, it is possible that the cells had three or more copies of the corresponding chromosomes before the BFB event, and at least two copies (from different haplotypes) were retained after BFB.

**Figure 7.**
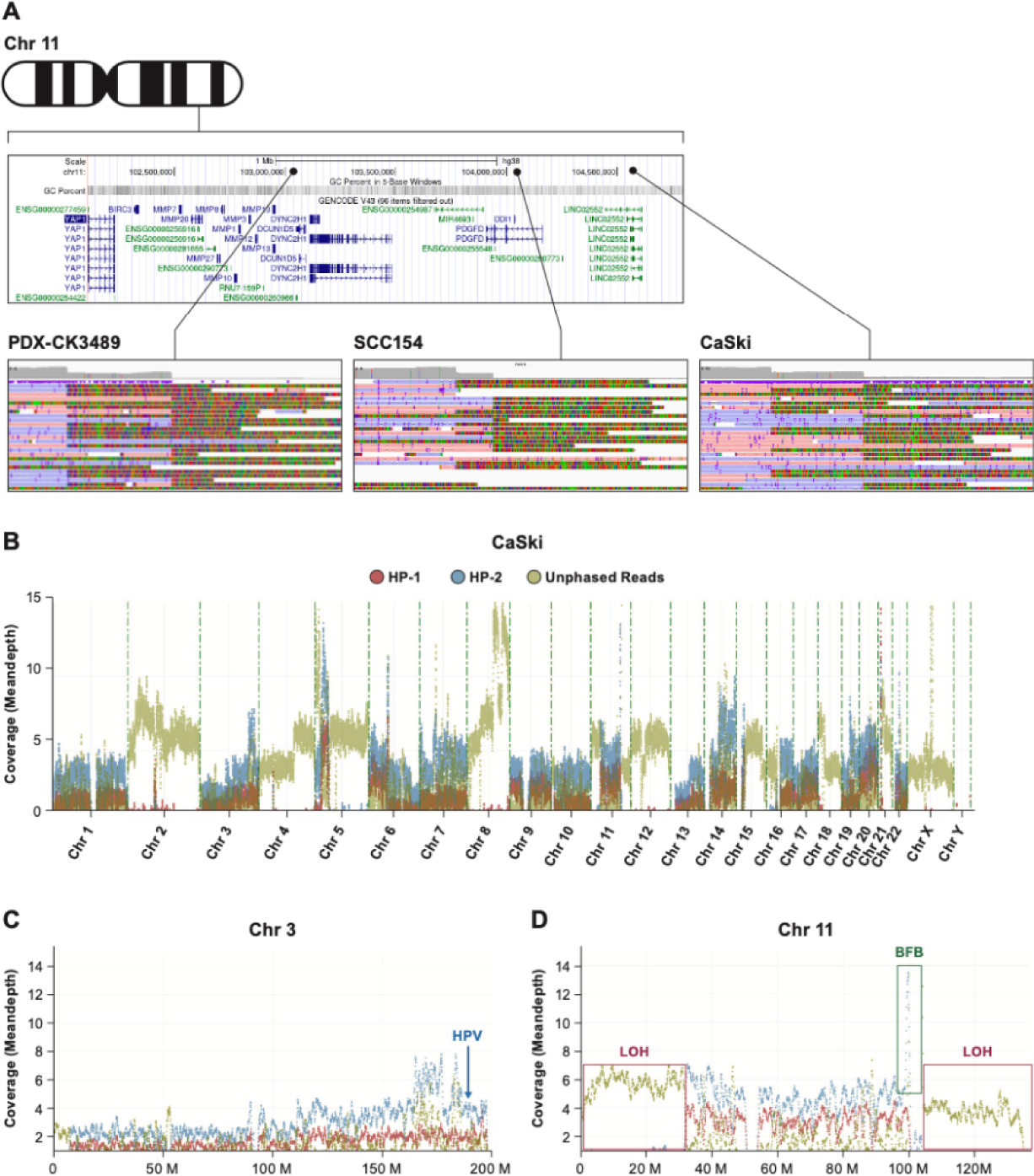
Map of the region of chromosome 11q22 with a cluster of 3 BFB events. **A.** Map of the chromosome 11q22 region containing the *YAP1*, *BIRC2*, and *BIRC3* genes is shown along with the location of three BFB events, in independent models. Detail of the read coverage aligned to HG38 is shown for the HNSCC PDX line (PDX-CK3489), SCC154 HNSCC cells and CaSki cervical cancer cells. All soft-clipped reads are inverted in relation to the aligned forward (blue) and reverse (pink) read segments. **B.** Phase reads copy number plot of the CaSki genome displaying reads assigned to haplotype 1 (HP-1) and 2 (HP-2) as well as unphased reads. Large blocks of unphased reads are due to loss-of-heterozygosity (LOH). **C.** Detail of chromosome 3 is shown indicating the location of multiple HPV integrations on 3q22-27 (131-188 MB). **D.** Plot of chromosome 11 showing the phased amplification of the *YAP1* region (BFB) and the deletion of one haplotype and LOH after the BFB event extending to the telomere.

We further attempted to localize the ends of derived chromosomes after BFB amplifications. We searched for reads containing telomere motifs and grouped them in 50 kb bins. This analysis identified telomeres at the expected locations (chromosome ends) but did not find any bins with more than three reads containing telomere sequences inside the BFB amplifications (**Supplemental Figure 4**). Analysis of clusters of read alignments with loose ends did not yield any rearrangements or abrupt sequence ends either (other than the reported foldback inversions and several focal events). This suggests that the derived chromosomes did not acquire canonical telomere sequences or fusion with another chromosome.

### *YAP1* amplification is caused by BFB events

Four cell lines (SCC154, CaSki, SNU-1000, and SNU-1299) have BFB events and amplify the *YAP1* oncogene and the *BIRC2* and *BIRC3* anti-apoptotic genes adjacent to each other on chromosome 11q22.1. Long-read WGS of a head and neck cancer (HNSCC) patient-derived xenograft (PDX) also revealed amplification of *YAP1*, *BIRC2,* and *BIRC3*, and copy number variant (CNV) loss of 11q distal to this region, and a Type I BFB event (**Figure 7A**). Analysis of phased reads in the CaSki genome revealed complex chromosome gains and losses across the genome consistent with data from short read WGS data (Akagi et al., 2014).

As YAP1 is a potent cervical cancer oncogene, and amplification of this locus is the most common oncogene amplification in cervical cancer, we examined additional genomic datasets. Analysis of the 293 cervical cancers in TCGA confirmed that 31 (11%) have *YAP1* amplification. **Figure 8A** shows the copy number values for genes proximal and distal to *YAP1*. In every tumor, the amplification occurs in a 3-4 MB region of Chr11q22 containing *YAP1*, *BIRC2,* and *BIRC3*. Distal to this region, the log2 CNV values are <1 for all genes on 11q (**Figure 8A**). This indicates that nearly all YAP1/BIRC2/BIRC amplifications in CESC are due to BFB events. We performed similar analyses for 20 cervical tumors in the ICGC-PCAWG dataset, 103 cervical SCC tumors in the MSKCC metastasis study, 90 Guatemalan cervical tumors (Lou et al., 2015)(Rossi et al., 2023), and 391 cervical SCCs in the AACR GENIE cohort (Consortium, 2017) (https://www.cbioportal.org/) (**Supplemental Figure 5**). In these studies, virtually all tumors with *YAP1* CNV gain show a focal gain of the *YAP1* region and loss of 11q22-tel. This result strongly suggests that deletion of Chr11q, followed by BFB cycles, leads to *YAP1*/*BIRC2*/*BIRC3* amplification.

**Figure 8.**
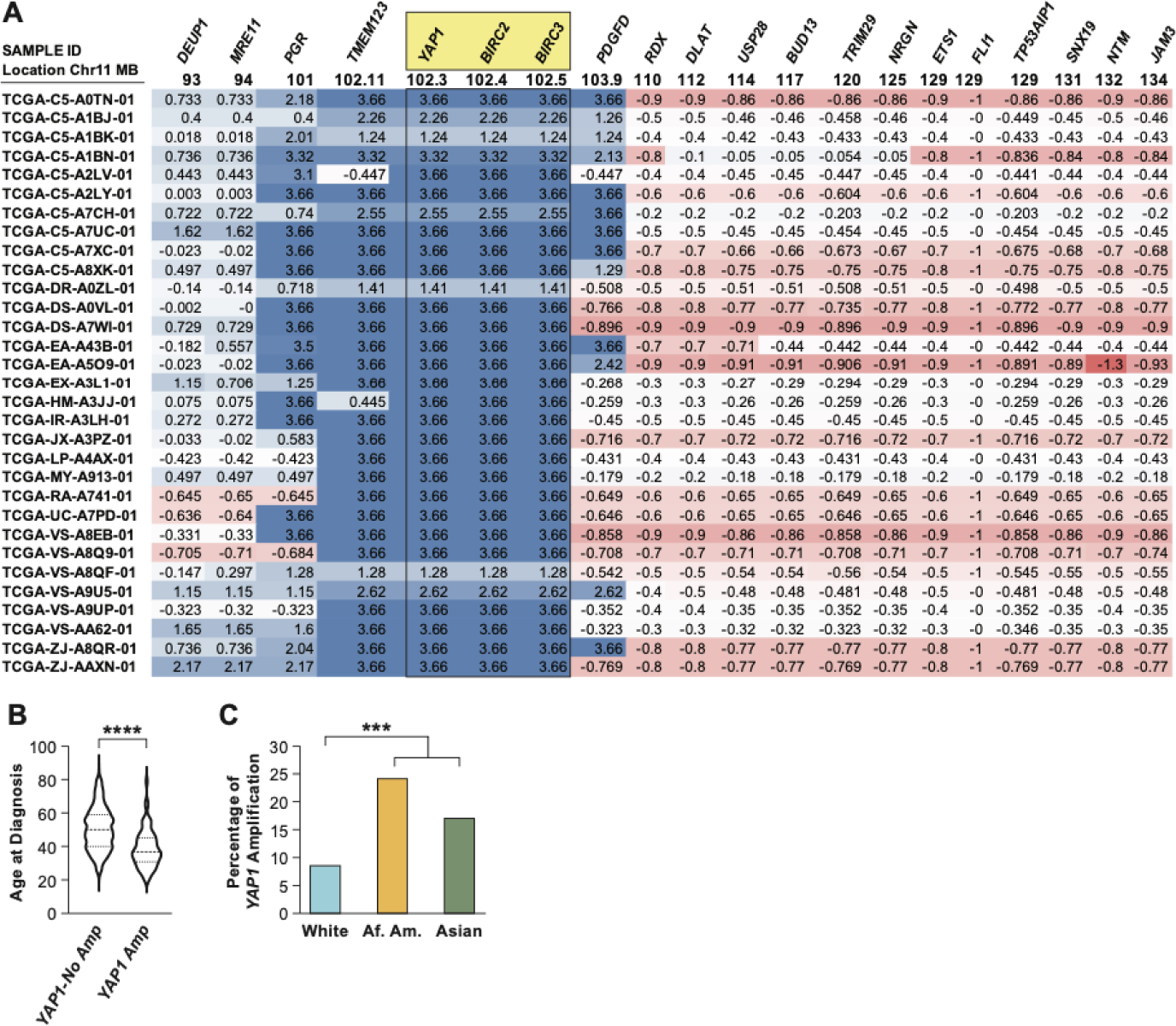
Analysis of *YAP1* gene amplification in 31 cervical tumors. **A.** Copy number data for genes centromeric and telomeric to *YAP1* in all TCGA cervical tumors with Log2 copy number values for *YAP1* >1. The location of the *YAP1*, *BIRC2* and *BIRC3* genes are highlighted. The location of each gene in MB is given below the gene symbol. **B.** Comparison of the age of diagnosis between women with YAP1-amplified and unamplified tumors **C.** Self-identified race of cervical cancer patients with and without *YAP1* amplification. Data from TCGA, MSKCC metastatic cancer, and AACR GENIE cohort were obtained from cbioportal (https://www.cbioportal.org) (Cerami et al., 2012; Gao et al., 2013).

Within TCGA, MSKCC, and GENIE, the frequency of *YAP1* gain in cervical cancer is the highest relative to all cancer types (**Supplemental Figure 6, Supplemental Table 5**). Interestingly, *YAP1* gain is also high in other cancer types caused by HPV, such as HNSCC, penile, anal, and vulvar cancers (**Supplemental Figure 6**). In addition, in these studies and the combined dataset, the median age of diagnosis of cervical cancer patients with *YAP1* amplification is only 36 years old, 14 years younger than patients without this CNV event. This data indicates that *YAP1* amplification leads to rapid cervical cancer development and progression. Finally, *YAP1* amplification is significantly (P=0.000x) more frequent in cervical tumors from African, Asian, and other women of non-European ancestry (**Figure 8C**). Therefore, targeted therapy for the cervical cancer subtype with an amplified *YAP1* region could help reduce cervical cancer health disparities.

## Methods

### Cell culture

The cell lines were obtained from the American Type Culture Collection (ATCC) and the Korean Cell-Line Bank and Cancer Research Center, Seoul National University (Ku et al., 1997). Cells were cultured in EMEM or RPMI-640 media with 10% Fetal Bovine Serum, 1% Pen Strep (10,000 units/mL of penicillin, 10,000 µg/mL of streptomycin, and 25 µg/mL of Amphotericin B) until 70-80% confluent. Cells were washed with 10 mL PBS and harvested using 2 mL trypsin per T-75 flask. All cell lines were confirmed by Identifier analysis, and regularly shown to be mycoplasma negative.

### PDX cell methods

The tissues for PDX-CK3489 were obtained from the NCI Patient-Derived Models Repository. The tissue was obtained from a 74-year-old HPV+ HNSCC male patient (collected on 05/2014). https://pdmdb.cancer.gov/web/apex/f?p=101:3:0::NO:3:P3_PATIENTSEQNBR:244. The frozen tissue (2-3 mm cube) was thawed on ice and washed with ice-cold PBS to remove DMSO (the component of the freezing media). We implanted the washed tissues under the skin of the right flank of the NSG mice. The animal was euthanized when tumors reached a maximum of 10% of body weight (∼2000 mm3). The tumor was harvested and frozen in 10% DMSO and 90% FBS freezing media until further use or analysis.

### DNA, and RNA extraction

Cells or tissues were washed with 10 mL PBS and harvested using 2 mL trypsin per T-75 flask. DNA was extracted and purified using a Gentra Puregene kit from Qiagen. Cell line RNA was prepared from 30 million cells using Trizol (ThermoFisher) and Poly-A+ RNA purified by DYNAL Dynabeads (Invitrogen). DNA was quantified by Nanodrop (Thermo Scientific) and Qubit (Thermo Scientific) and stored at 4-L; RNA was quantified by Qubit and stored at −80-L.

### PCR Amplicon and Sanger sequencing validation

The Primer3 online program (Untergasser et al., 2012) was used to design primers for PCR amplification. Overlapping primers were designed to span the entire 7.9 kb HPV genomes. Cell line DNA was amplified using a long-range PCR kit from New England BioLabs. The samples were run on a 1.5% agarose gel at 100V for 1 hour and imaged with a BioRad ChemiDoc Image System. To determine the HPV lineage and sublineage, we purified the PCR products and sequenced them on an Applied Biosystems® 3500xL Genetic Analyzer from Thermofisher Scientific. We analyzed the sequences were using DNASTAR SeqMan Ultra and SeqBuilder Pro software.

### HLA typing by DNA sequencing

HLA typing was performed using targeted next-generation sequencing (NGS) with locus-specific primers used to amplify a total of 26 polymorphic exons of HLA-A & B (exons 1 to 4), C (exons 1 to 5), E (exon 3), DPA1 (exon 2), DPB1 (exons 2 to 4), DQA1 (exon 1 to 3), DQB1 (exons 2 & 3), DRB1 (exons 2 & 3), and DRB3/4/5 (exon 2) genes with Fluidigm Access Array (Fluidigm Corporation, South San Francisco, CA 94080 USA). The 26 Fluidigm PCR amplicons were pooled and subjected to sequencing on an Illumina MiSeq sequencer (Illumina, San Diego, CA 92122 USA). HLA alleles and genotypes are called using the Omixon HLA Explore (version 2.0.0) software (Omixon Biocomputing Ltd., Budapest, Hungary).

### Long-read DNA Sequencing

We used the Ligation Sequencing kit (SQK-LSK109, SQK-LSK110, Oxford Nanopore Technologies) to perform long-read DNA sequencing of the cell line DNA samples. We used either unsheared DNA or DNA that was sheared to 8-20 kb with a G-tube (Covaris) from 1-4 ug of input DNA. We applied adaptive sampling to selected samples using a reference FASTA file containing the human HG38 genome and high-risk HPV type FASTA file and a BED file that specified the regions of interest, such as cancer genes, integration loci, and HPV sequences. We loaded the samples onto MinION R9.4 flow cells on a GridION instrument and performed the sequencing runs. **Supplemental Table 1** provides the details of each sequencing run, and **Supplemental Table 2** summarizes the WGS coverage of each cell line.

### Identification and characterization of HPV integration events

WGS reads from ligation sequencing or adaptive sampling were mapped to the hg38 human genome assembly and a file containing 13 hrHPV types. Reads aligning to both human and HPV were identified, and individual segments of human and HPV DNA were identified with BLAT and BLAST. We validated most of the junctions between human and HPV DNA by PCR and Sanger sequence, and we generated the final assemblies of the integrated HPV genomes using SeqMan Ultra software.

### Identification of Somatic mutations

For cell lines sequenced by the COSMIC cell line or CCLE projects, driver mutations in the 299 PanCancer Driver genes were identified from public data. We confirmed most of these mutations in our ONT data by manually reviewing BAM files. In previously uncharacterized cell lines, mutation hotspots and selected genes were manually reviewed in IGV to identify mutations. Mutations in the *PIK3CA* exons 9 and 20 were confirmed by PCR and Sanger sequence.

### Analysis of BFB events in cancer cohorts

The TCGA, AACR Genie, and Memorial Sloan Kettering datasets were accessed through cbioportal (http://www.cbioportal.org/). Copy number (log2) values of >2 for YAP1 and <0 for 11q telomeric genes were identified as potential BFB events. Data for age at diagnosis and race/ethnicity was obtained from the same site. Whole-genome haplotype-specific coverage plots were generated using Wakhan (https://github.com/KolmogorovLab/Wakhan; manuscript in preparation).

### Full-length RNA sequencing

Cell line RNA (500ng Poly-A+) was sequenced using the Direct RNA kit (DCS109) and Direct RNA sequencing kit (SQK-RNA002, Oxford Nanopore Technologies). RNA libraries were loaded onto MinION R9.4 flow cells on a GridION instrument (Oxford Nanopore Technologies). Transcriptome fastq files were aligned and reads assigned to genes using the Exome workflow in EPI2ME (<u>EPI2ME™ :: Dashboard</u> (nanoporetech.com)). Reads were normalized as reads per million (RPM). HPV reads were identified by alignment to a file with 13 high-risk HPV sequencing plus HPV26 and HPV30 using EPI2ME. HPV reads were counted and normalized as RPM and E6 splicing was assessed by manual counting of spliced and unspliced reads in BAM file produced in the CGC platform (https://cgc.sbgenomics.com/).

### Detection of inverted reads and putative BFB Events

The Breakage-Fusion-Bridge events were detected using Severus (https://github.com/KolmogorovLab/Severus; manuscript in preparation). Briefly, clusters of split read alignments were used for each cell line to identify breakpoints. Then clusters with foldback inversions and change in coverage were selected as BFB candidates. Identified breakpoints were further manually investigated using IGV.

### Validation of inverted sequences

Inverted reads at BFB sites were confirmed by designing primers specific to single-copy sequences at the junctions using the Primer3 tool. We performed single primer PCR reactions with the forward primers and resolved the products on agarose gels. We selected the primers that produced bands only in the cell line with the BFB event and added barcodes to them. We sequenced the barcoded products on ONT Flongle flow cells and aligned the sequences to the HG38 human genome assembly. We mapped the reads to confirm the presence of BFB events.

### Bioinformatics and statistics

We performed sequence alignment of the FASTQ files obtained from the sequencing runs to the human genome (HG38) or HPV genomes using the EPI2ME server (https://epi2me.nanoporetech.com). We used the Fastq Human Alignment GRCh38 app for human genome alignment and the Fastq Custom Alignment workflow for HPV genome alignment, providing a FASTA file of HPV16 or HPV18 or 13 other high-risk HPV types as a reference. We combined the alignment data for HG38 and HPV with Excel or in Filemaker (Claris) and added read length and adaptive sampling decision data, if applicable. Reads of interest were manually extracted and mapped using BLAT (Kent, 2002) (https://genome.ucsc.edu) or BLAST. BAM files produced by EPI2ME were merged and indexed using the BamTools Merge and SAMtools Index tools in the Cancer Genomics Cloud (CGC) (Lau et al., 2017). We also ran an Oxford Nanopore Technologies WGS Data Processing pipeline on CGC, based on a Broad Institute pipeline, to align human and HPV reads and produce BAM files. BAM files were viewed in the Integrated Genome Viewer (IGV) (Thorvaldsdottir et al., 2013).

### Pipelines

DNA reads were analyzed using fast5 raw data by base-calling using Guppy/4.5.4 [8] (https://nanoporetech.com/). Modified base-calling was performed using Megalodon/2.3.3 https://github.com/nanoporetech/megalodon. Structural variation calling was carried out with the Nanopore pipeline-structural-variation with modifications. The entire workflow is available at https://github.com/NCI-CGR/Nanopore_DNA-seq. RNA reads were processed using the fast5 raw data and base-called using BINITO /v0.3.7 and aligned to the HG38 genome using Minimap2/2.17. Isoforms were detected and quantified using Stringtie2/2.1.5 and Freddie (https://github.com/vpc-ccg/freddie). The Freddie program calls the Gurobi package (www.gurobi.com) to solve optimization problems. The entire workflow is available at https://github.com/NCI-CGR/Nanopore_RNA-seq.

### HPV lineage classification

HPV lineage and sublineage was determined using MEGA (Kumar et al., 2018) and ClustalOmega. Cell line FASTA files were exported from the GridION and translated into amino acid sequences using MEGA. Maximum likelihood phylogenetic trees were constructed to determine the closest HPV variant match for each cell line with Bootstrap values of >80 was used. Domain sites for HPV genes E6, E7, E1, E2, L1, and L2 were set to look for amino acid changes in each sequence specified by that region. Alignments were confirmed using Clustal Omega.

### Statistical analyses

Statistical analysis was performed in GraphPad. We used Fisher’s exact test to assess the relationship between HLA homozygous and heterozygous for type A, B, C. We used an unpaired t-test with Welch’s correction to ascertain the relationship between age of onset of *YAP1* amplified tumors. In all calculations, a p-value of 0.05 or less was deemed significant.

## Discussion

We performed a multi-omics analysis of the cervical and head and neck cancer cell lines, including complete HPV sequencing, HPV integration analysis, copy number alterations, HLA gene sequencing, and human and HPV gene expression. The main goal of this project was to characterize a panel of HPV+ cervical and head and neck cancer cell lines using long-read methodology to further understand HPV integration events, and genomic rearrangements. Cervical cancer is under-represented in major international cell line collections such as the NCI-60 panel (0 cervical lines), COSMIC (19 lines), and CCLE (14 lines) (**Supplemental Table 4**). Our panel of 22 cell lines includes 18 cervical cancer cell lines, 10 of which are not represented in any current cell line panel (**Supplemental Table 4**); includes the major cervical cancer (CESC) histological types (squamous cell carcinoma, adenocarcinoma and small cell carcinoma) as well as a comparison group of HPV+ HNSCC cell lines. The panel includes cell lines from individuals of European Asian, and African American origin. We document the diversity of HPV types, HPV sublineages, and integration loci. Long-read DNA sequencing with Oxford Nanopore platforms was used for long-range characterization of the HPV integration loci. Finally, full-length RNA sequencing was performed using direct cDNA and direct RNA approaches for transcriptome analysis.

Cervical cancer disproportionally affects women living in poverty, especially in Sub-Saharan Africa, where it causes about 50,000 deaths every year (Mboumba Bouassa et al., 2017). African American women also face a higher risk of cervical cancer mortality, with a two-fold increase in age-adjusted death rates compared to other ethnic groups in the United States. We found that *YAP1*/*BIRC2*/*BIRC3* gene amplification, which is associated with earlier onset of cervical cancer, is three times more prevalent in cases of cervical cancer in African American women. This finding suggests that this genomic alteration (*YAP1*/*BIRC2*/*BIRC3* gene amplification) contributes to the ethnic disparity in cervical cancer mortality. Cohen et al. document that age-adjusted mortality from CESC-SCC is significantly higher (1.9-fold) in African Americans in the U.S. (Cohen et al., 2023). *YAP1* overexpression in cervical cells activates EGFR and its ligands, which are potential therapeutic targets for EGFR inhibitors (He et al., 2015). In addition, YAP1-targeted agents are currently being tested in clinical trials (Liu et al., 2019).

HPV infection and integration can cause genome instability in HPV-driven cancers, leading to genome rearrangements and the formation of extrachromosomal HPV/human hybrids (Keiko Akagi, 2022). However, little is known about other copy number alterations in cervical cancer. In this study, we used long-read sequencing to analyze cancer cell lines. A single BFB event was observed in six cell lines, and BFB events on more than one chromosome were identified in three cell lines. Although BFB events are common mechanism of genome instability in cancer cell genomes, few studies have characterized the sequence features of the inversion sites involved in BFB cycles. We found that the breakage ends underwent exonuclease digestion and that the fusion sites showed DNA insertions and rearrangements. We categorized three types of Breakage-Fusion-Bridge (BFB) events: Type I (with no insertions or rearrangements); II (with insertion from another chromosome), and III (with additional local rearrangement). Long read sequencing was crucial in determining the precise structure of the insertions and rearrangements at BFB junctions. Five cell lines showed *YAP1* amplification associated with BFB, making this one of the most common driver events in the cell line panel. In addition, amplification of the *YAP1*/*BIRC2*/*BIRC3* loci was one of the most frequent copy number alterations in cervical tumors in the TCGA, MSKCC metastatic cancer, and AACR GENIE cohorts (**Supplemental Figure 7**) (Cancer Genome Atlas Research et al., 2017; Consortium, 2017). Furthermore, we found genes inactivated by BFB, including the *WT1* tumor suppressor gene. Therefore, BFB events are essential in the progression of cervical cancer through the activating or inactivating of essential cancer driver genes.

*YAP1* is an oncogene that is amplified in 15% of CESC cases, and overexpression of *YAP1* in mouse cervical cells can cause cervical carcinogenesis without HPV infection (Nishio et al., 2020). Furthermore, *YAP1* expression repressed differentiation of HPV-infected basal cells in the cervix (Hatterschide et al., 2022). Our data strongly suggests that nearly all *YAP1* amplification is the consequence of BFB events. We found that *YAP1* amplification is associated with a 10-14 year earlier age of diagnosis of cervical cancer. In addition, previous IHC studies showed that the YAP1 protein is high in both high-grade pre-cancer and cervical cancer (Hatterschide et al., 2022). This result suggests that *YAP1* amplification is both an early event in cervical cancer and leads to the rapid progression of the disease. Furthermore, we consistently found *BIRC2* and *BIRC3* amplified along with *YAP1*, demonstrating that these genes also play a crucial role in cancer progression. High levels of *BIRC2* and *BIRC3* could contribute to cervical cancer progression by conferring resistance to T-cell-mediated cell death.

Our analysis of the telomeric side of BFB amplifications suggests that the derived chromosome did not acquire a canonical telomere sequence, nor was it fused with another chromosome. Therefore, the exact mechanisms of stabilizing the derived chromosome after BFB remain unclear. It has been hypothesized that the chromosome may remain in a double-strand break state after BFB, and our observations are consistent with this explanation (Dewhurst et al., 2021). We expect that additional long-read sequencing of diverse tumors at higher depth will provide further information on this critical class of chromosome alteration.

HPV infections and early pre-cancerous lesions are often rapidly cleared, indicating a robust immune response to virally infected cells (Schiffman et al., 2016). Furthermore, genome-wide association studies of cervical cancer reveal strong associations with the MHC on Chr 6p, principally to HLA class II and HLA class I genes (Leo et al., 2017) and HLA class I genes are frequently mutated in CESC (Dean et al., in preparation). We found that 56-61% of 19 CESC cell lines are homozygous across the HLA class I and II genes, significantly higher than the homozygosity in cell lines of other cancer types, supporting a key role for the MHC in cervical cancer progression.

Currently, there is no targeted therapy for cervical cancer. However, *PIK3CA* is the most common oncogene driver mutation, which is present in six out of 21 of our cell lines. BYL719/Alpelisib is a specific PI3K inhibitor approved for treating a subset of breast cancers (Andre et al., 2019). Preclinical data show that BYL719 inhibits cell lines with *PIK3CA* mutations (Lou et al. manuscript in preparation). As described above, *YAP1* amplification is also frequent in cervical cancer and is largely mutually exclusive of *PIK3CA* mutation (Lou et al. manuscript in preparation). Therefore, *YAP1* amplified tumors represent a second cervical cancer subtype potentially amenable to targeted therapy. Our cell panel provides model systems to study targeted inhibitors and immune therapies and advance the therapeutic options for cervical cancer.

Our cell lines confirm the distribution of HPV integration across many human chromosomes, but 80% of integrations are in known HPV integration hotspots or near super-enhancers (Warburton et al., 2021). However, most cell lines have integration in or near a gene of potential oncogenic importance and activate the expression of at least one gene at the locus. As HPV brings two potent oncogenes to the cell, driver gene mutations are less prevalent in cervical cancer. In our cell lines, there was an increase in mutations in cancer driver genes in cell lines without HPV or with unknown-risk HPV types. Mutations in *TP53, RB1,* and other cancer driver genes, may substitute for the absence or low activity forms of the E6/E7 oncogenes. For example, the cell line C33A is HPV-negative (we confirmed this by long-read sequencing) and is homozygous for pathogenic mutations in both the *TP53* and *RB1* genes. HT-3 has an integrated copy of HPV30, an unknown-risk type, and has mutations in *RB1* and *TP53* that may potentiate the E6 and E7 proteins of HPV30.

Our study has several limitations, such as the under-representation of cell lines derived from adenocarcinomas, an aggressive subtype of cervical cancer, and only one cell line (HeLa) from a woman of African origin. There are likely unknown biases in what classes of cervical tumors were established as cell lines. Episomal-only tumors and cell lines are known to be unstable, and only SNU-1000 retains extrachromosomal HPV16. We described only 19 CESC cell lines, but 10 are unique to this panel (**Supplemental Table 4**). For several cell lines, limited coverage did not allow the detection of SNVs genome-wide or the identification of all chromosome alterations, including BFB events.

In summary, using a panel of 22 cervical and HPV-positive cell lines and long-read sequencing, we comprehensively characterized the structure of HPV integrations and the consequences on gene expression. We show that HPV-negative and non-high-risk HPV type cell lines have more driver mutations, including in *TP53* and *RB1,* to compensate for the low or absent activity of HPV E6 and E7. In addition, we characterized the sequences at the inversion junctions of breakage-fusion-bridge events and provided new insight into the formation of these critical chromosome rearrangements in cancer. These cell lines can serve as models for specialized treatments of cervical cancer.

## Data Availability

All data produced are available online at https://www.ncbi.nlm.nih.gov/bioproject/PRJNA772772

## Acknowledgements

This project has been funded in whole or in part with federal funds from the Frederick National Laboratory for Cancer Research, under Contract No. 75N91019D00024, and the NIH Intramural Program, Frederick National Lab, and the Center for Cancer Research. The content of this publication does not necessarily reflect the views or policies of the Department of Health and Human Services, nor does mention of trade names, commercial products, or organizations imply endorsement by the U.S. Government. This Research was supported in part by the Intramural Research Program of the NIH, Frederick National Lab, Center for Cancer Research. The authors would like to acknowledge the American Association for Cancer Research and its financial and material support in the development of the AACR Project GENIE registry, as well as members of the consortium for their commitment to data sharing. Interpretations are the responsibility of study authors. The Seven Bridges Cancer Research Data Commons Cloud Resource has been funded in whole or in part with Federal funds from the National Cancer Institute, National Institutes of Health, Contract No. HHSN261201400008C and ID/IQ Agreement No. 17X146 under Contract No. HHSN261201500003I and 75N91019D00024. Daria A. Gaykalova was supported by a Research Scholarship Grant, RSG-21-020-01-MPC from the American Cancer Society, and by R01DE027809 from the National Institute of Health.

## Notes

### Competing Interest Statement

The authors have declared no competing interest.

