## Supplemental Figures for "Insights into the Mechanisms and Structure of Breakage-Fusion-Bridge Cycles in Cervical Cancer using Long-Read Sequencing"

**Supplemental Figures with Legends**

**Supplemental Figure 1**

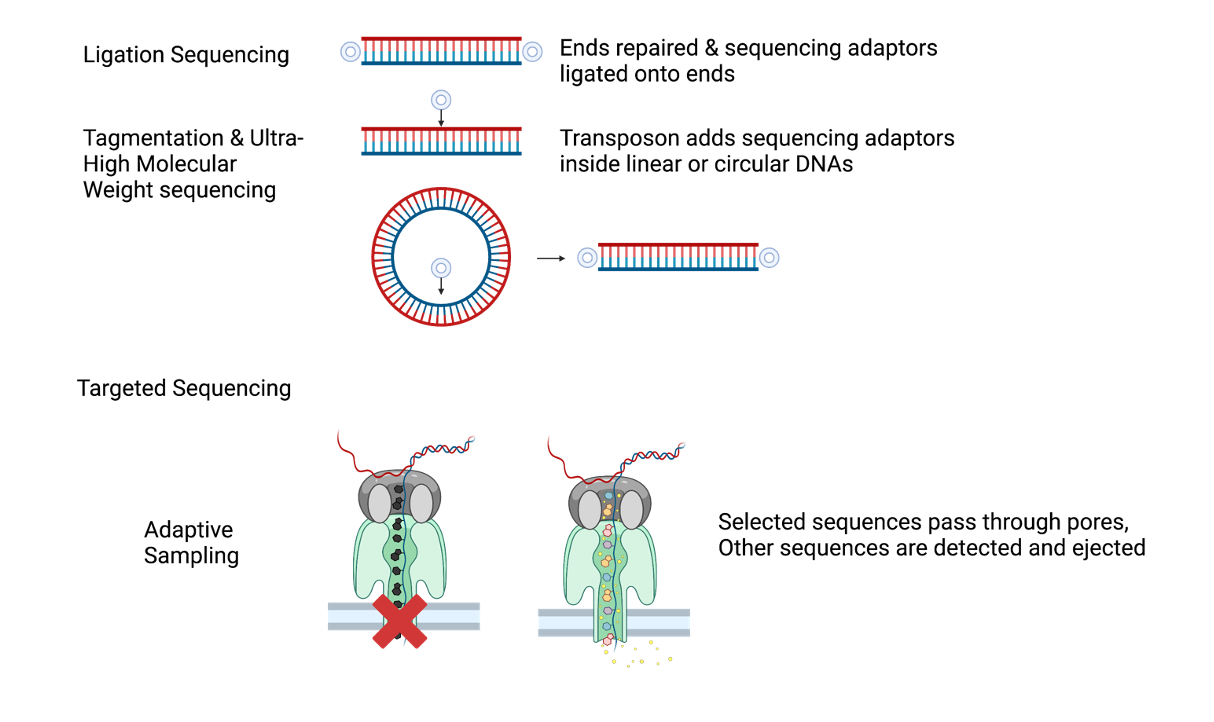

**Figure S1. Model of the long-read whole genome sequencing (WGS) sequencing strategy.** Libraries were prepared using Ligation Sequencing in which sequencing adaptors are ligated onto linear DNA molecules. Tagmentation is another option not used in this study. In selected cases adaptive sampling was used to enrich for cancer-associated genes, integration sites, and HPV.

Supplemental Figure 2
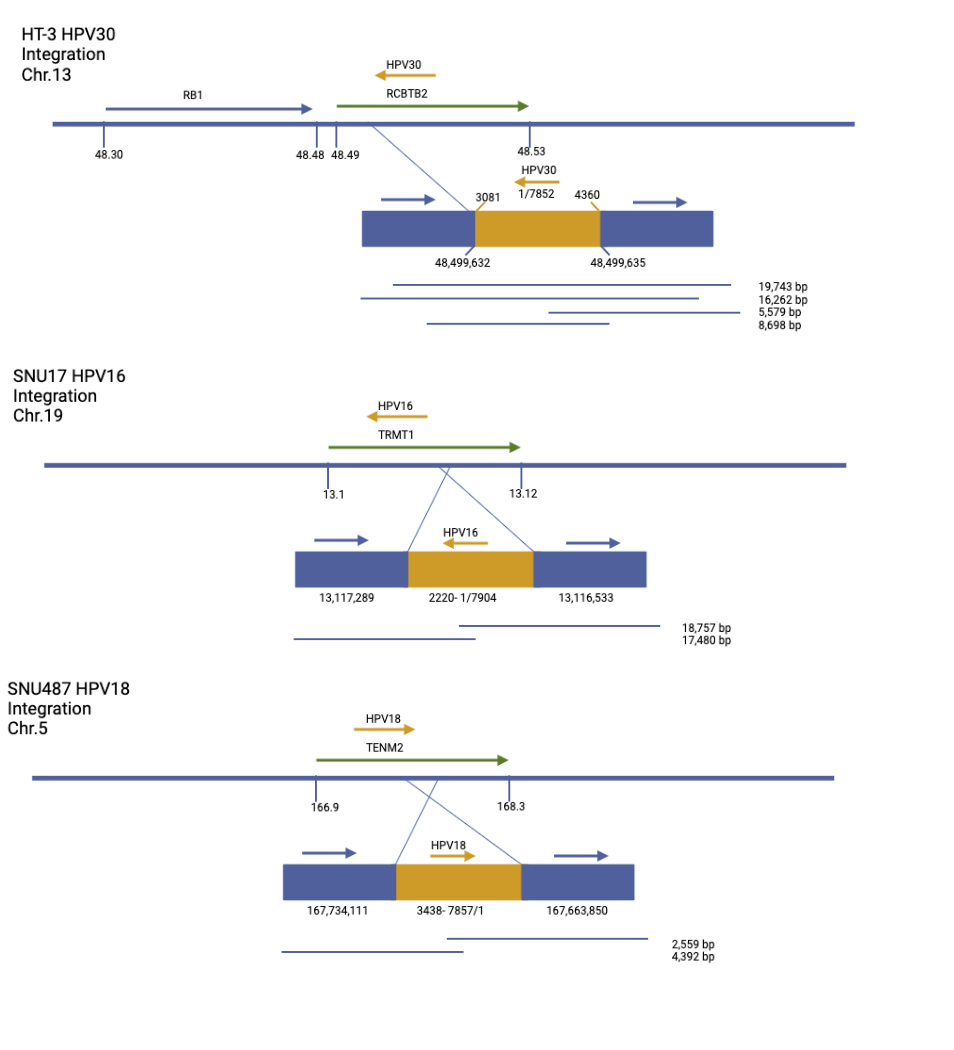

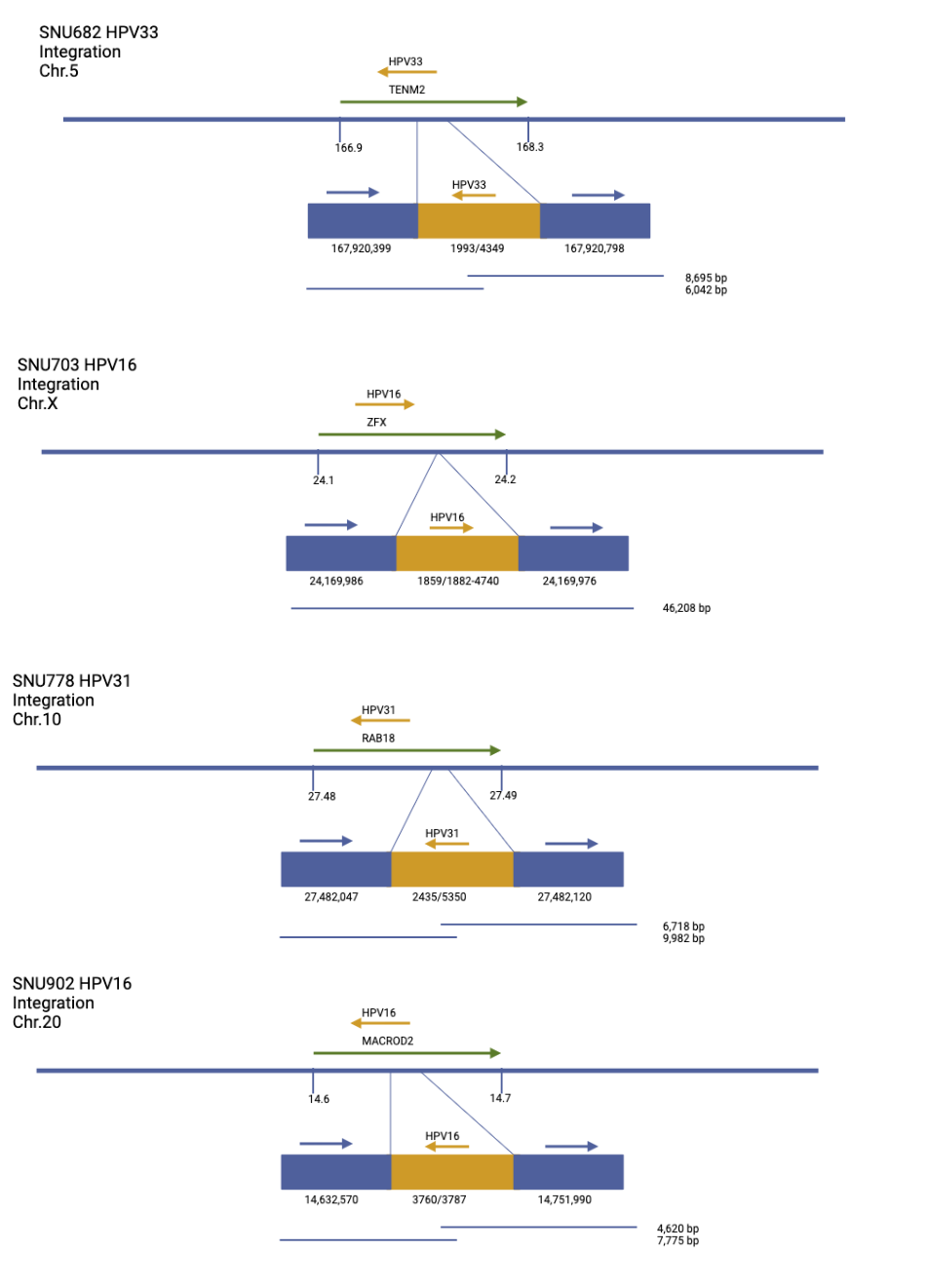

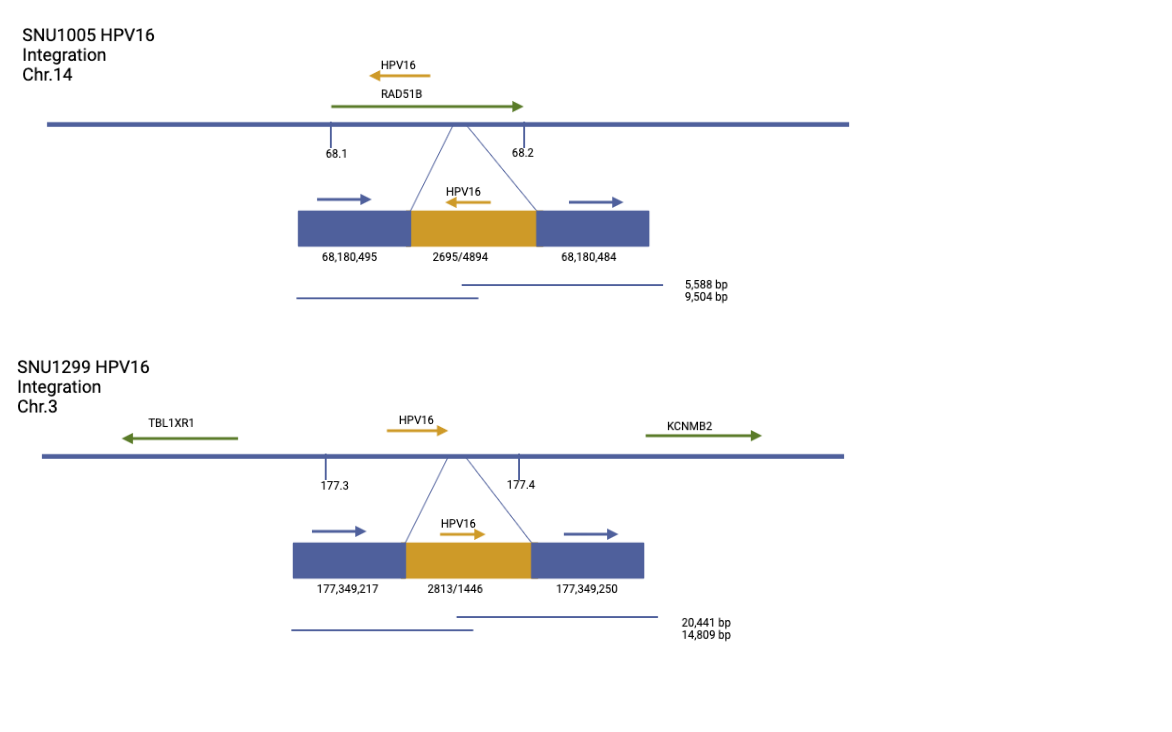

**Figure S2. Newly described HPV integration loci.** **A-I**, The location of integration into the human genome. Green arrows indicate genes found on the chromosome at or near the integration site, yellow arrows indicate the direction, type, and integration point of the HPV in that specific cell line. Blue blocks represent the human genome while yellow block represents HPV sequences.

Numbers below blue blocks indicate the position of integration, numbers below yellow block shows the position of the HPV virus integration. Blue lines show reads obtained by ONT across the integration site with respective size of reads.

Supplemental Figure 3

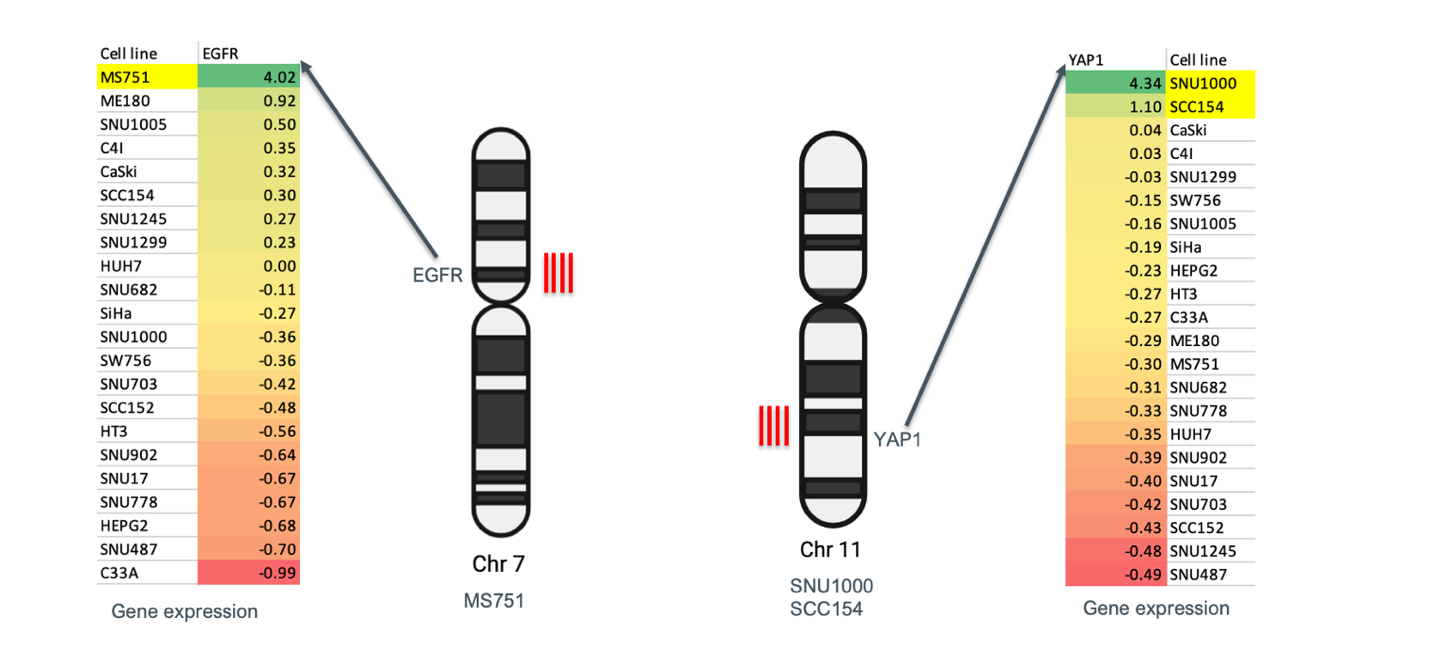

**Figure S3. Cell lines with oncogene amplification.** The normalized gene expression values for *EGFR* and *YAP* with highest expression at the top in green and lowest expression on the bottom in red.

Supplemental Figure 4

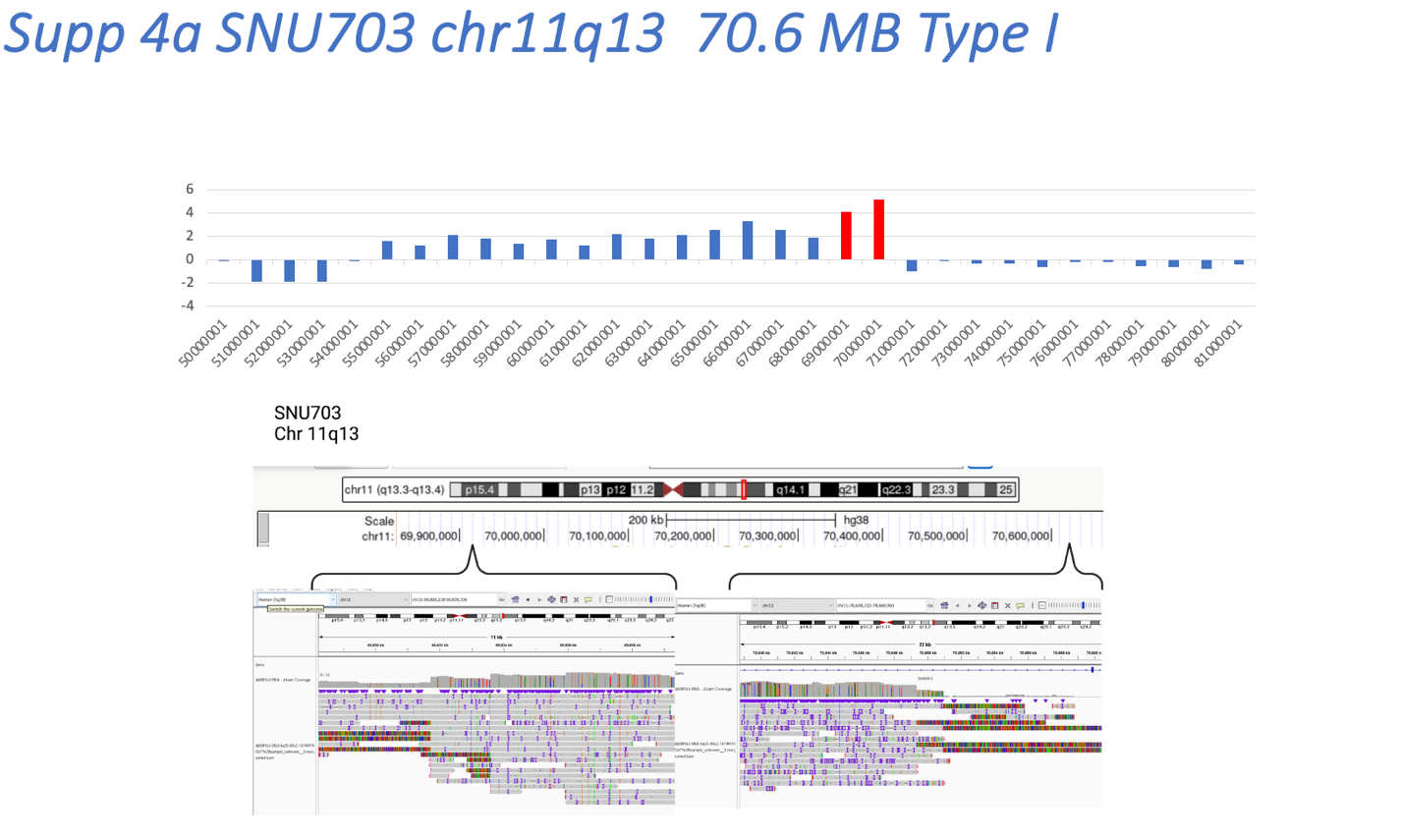

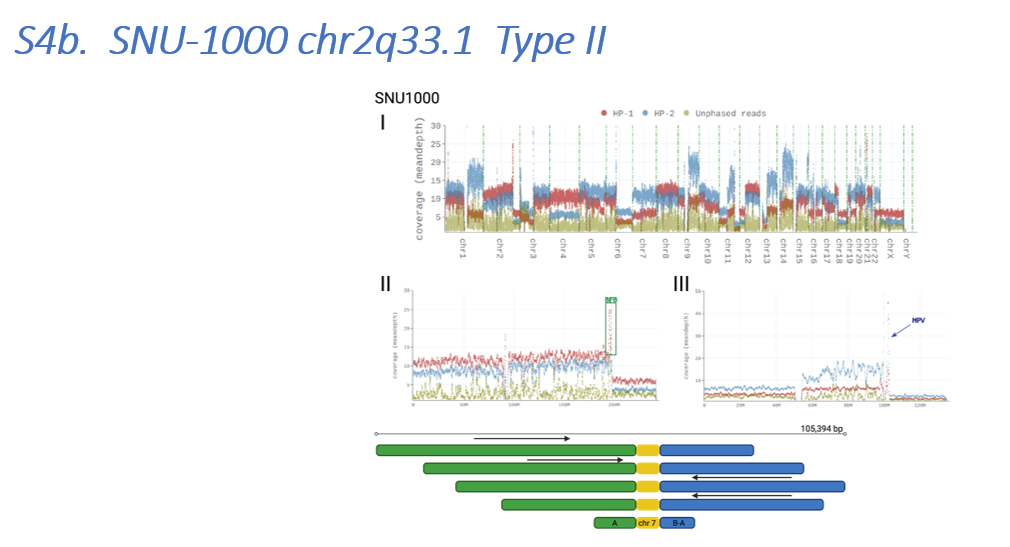

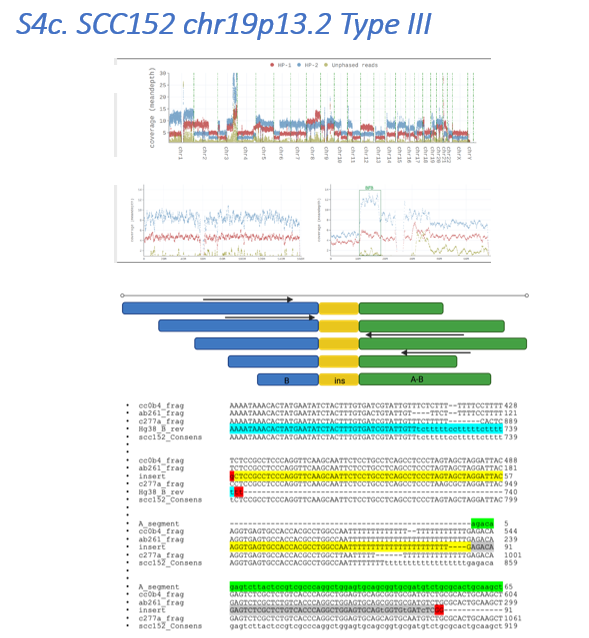

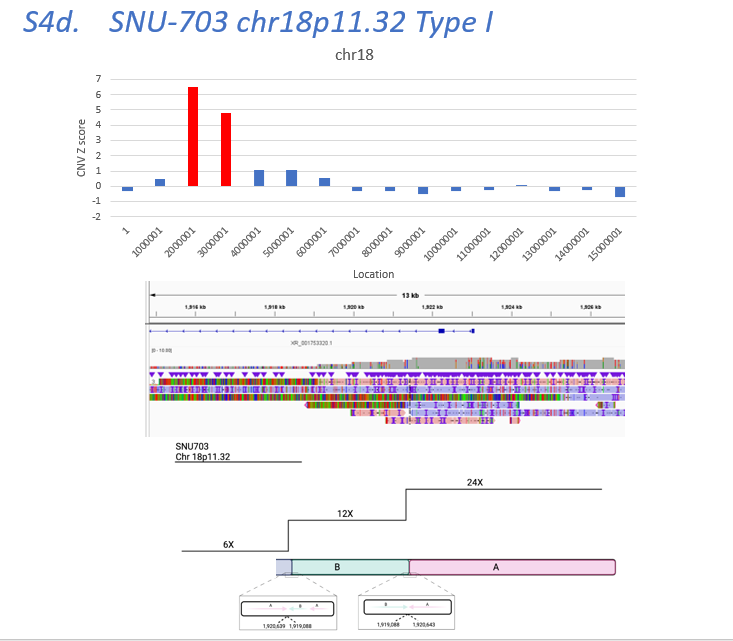

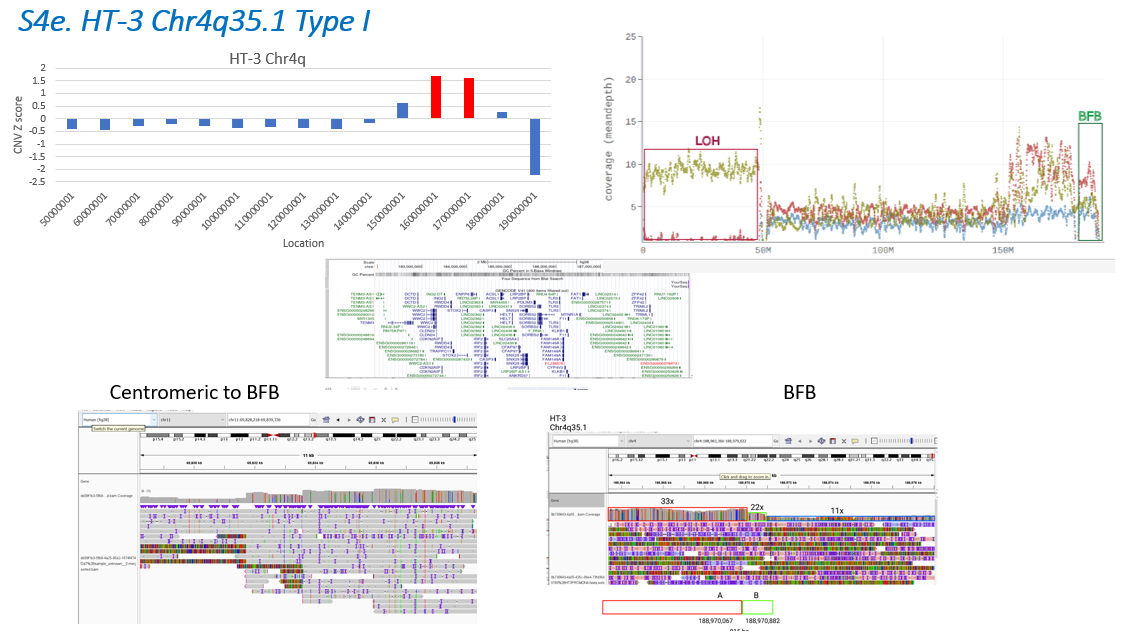

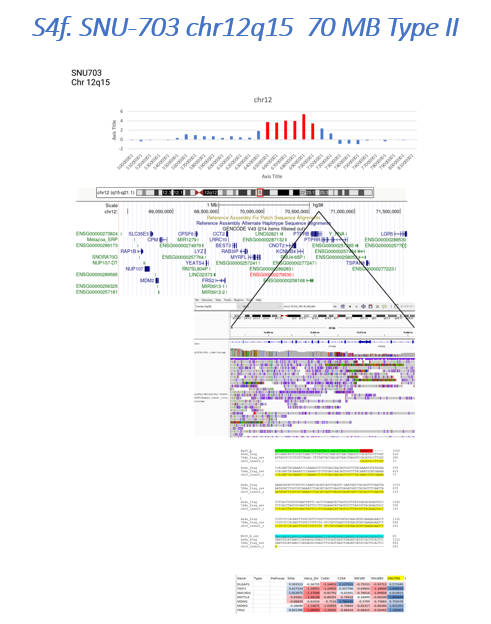

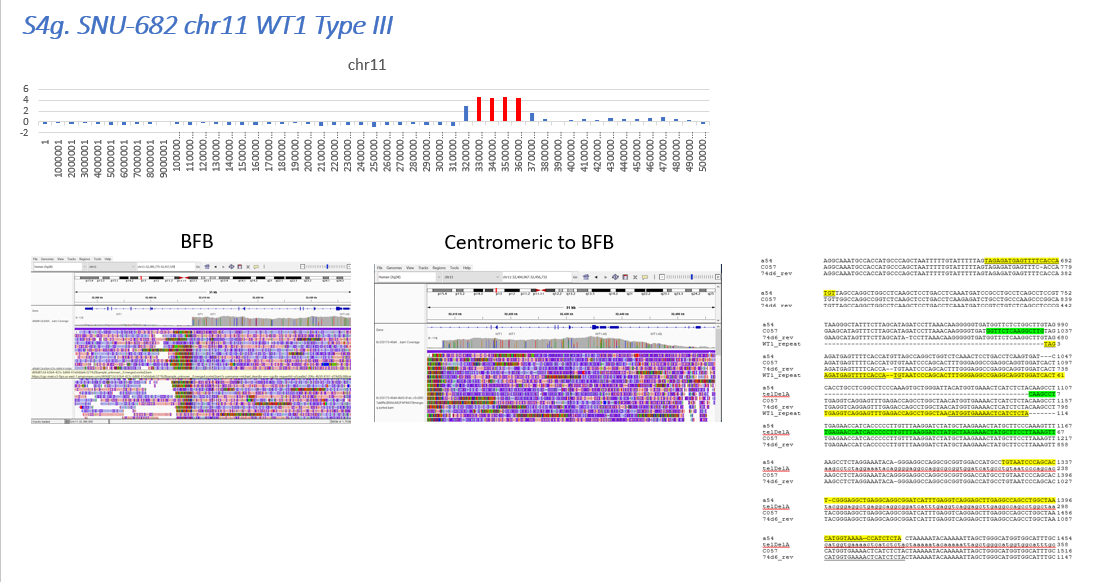

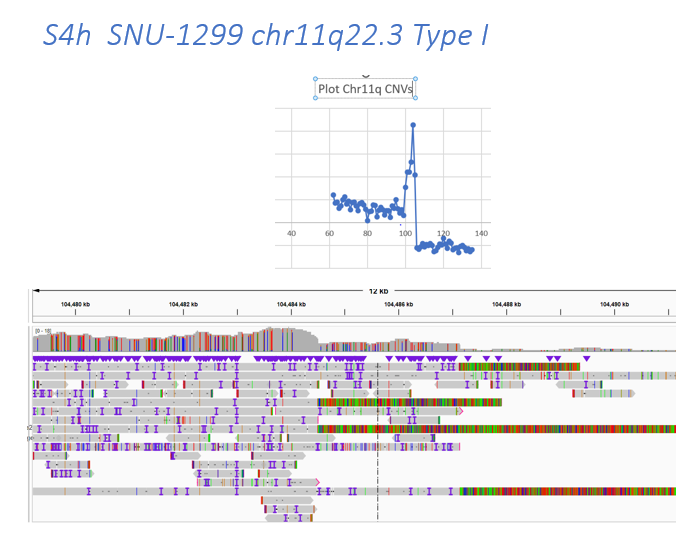

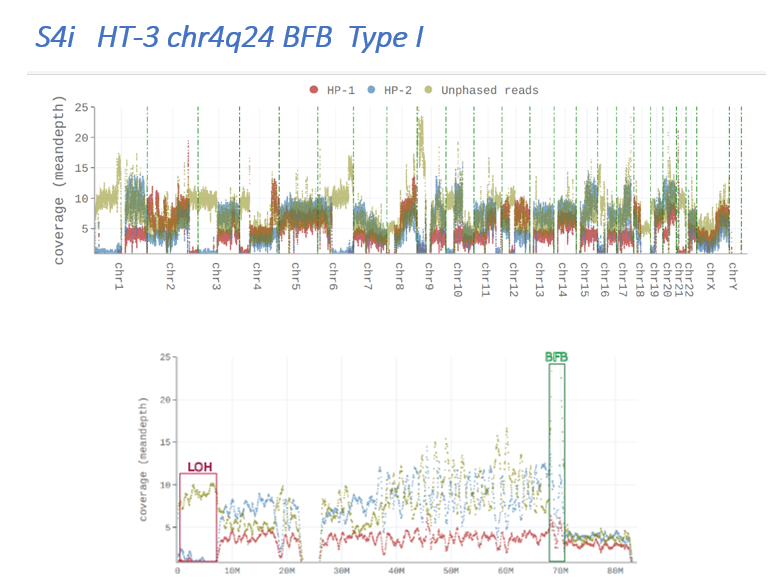

**Figure S4. Copy number plots and sequences at BFB sites. A,** The SNU-703 cell line showing a BFB region on chr11q13. A copy number plot of chr 11 is shown with amplified segments in red. Sequence detail of the BFB junction is shown below on the left, and on the right is a region where inverted reads are seen on the centromeric side. **B**, a phased whole genome plot of SNU-1000 (I) is shown along with the BFB site on chr 2 (II) and the HPV integration and *YAP1* amplification site on chr 11 (III). Below is a diagram of reads at the chr 2 BFB site showing the segment of chr7 (in yellow) inserted at the junction, along with the sequence of this region in individual ONT reads. **C,** Alignment of chr19 sequences at the BFB junction in SCC152. A phased WGS plot is shown with details of the BFB region on chr 19. The diagram below shows B segments (Blue) and A-B segments (Green) with inserted sequences from chr19:11133949-11134055 (yellow). An alignment of the corresponding sequences is below the diagram. **D,** details of SNU-703 BFB in chr18. A copy number plot of chr18 is shown along with aligned reads and a diagram of the structure of the junction at the breakage site. **E,** For HT-3 a copy number plot and phased plot of chr 4 is shown indicating the BFB site and a region of LOH on 4p. Aligned reads centromeric (left) and at the BFB site (right) are shown along with the coordinates of segment A and B. **F,** Details of the chr12 BFB event in SNU-703 cells. A copy number plot is shown along with the genes in the region, aligned reads and the details of the inserted segment from chr 8 (Yellow). At eh bottom is a Z-score plot of gene expression showing high expression of the methylase *METTL4* and several other genes in that region of chr 12q15. **G,** a copy number plot of chr11 is shown for the SNU-682 cell line along with read alignments of the BFB region and the region centromeric showing a gradual decline in copy number. Alignment of sequences at chr11 *WT1* junction. Green are segment A sequences. At the right are aligned sequences showing an insertion (yellow) of sequences that are locally rearranged sequences and inserted at the junction/inversion site. Bold and underlined sequences are palindromic. **H,** a copy number plot of chr 11q in SNU-1299 cells is shown above aligned reads at the BFB junction. **I**, A phased read plot of the HT-3 genome along with the details of chr4 showing that the BFB impacts only haplotype 2 (HP-2).

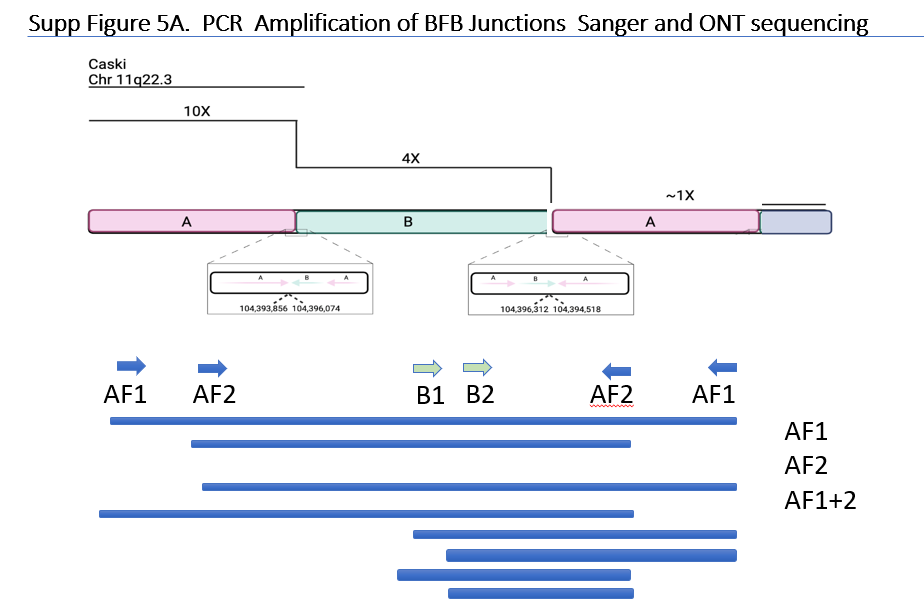

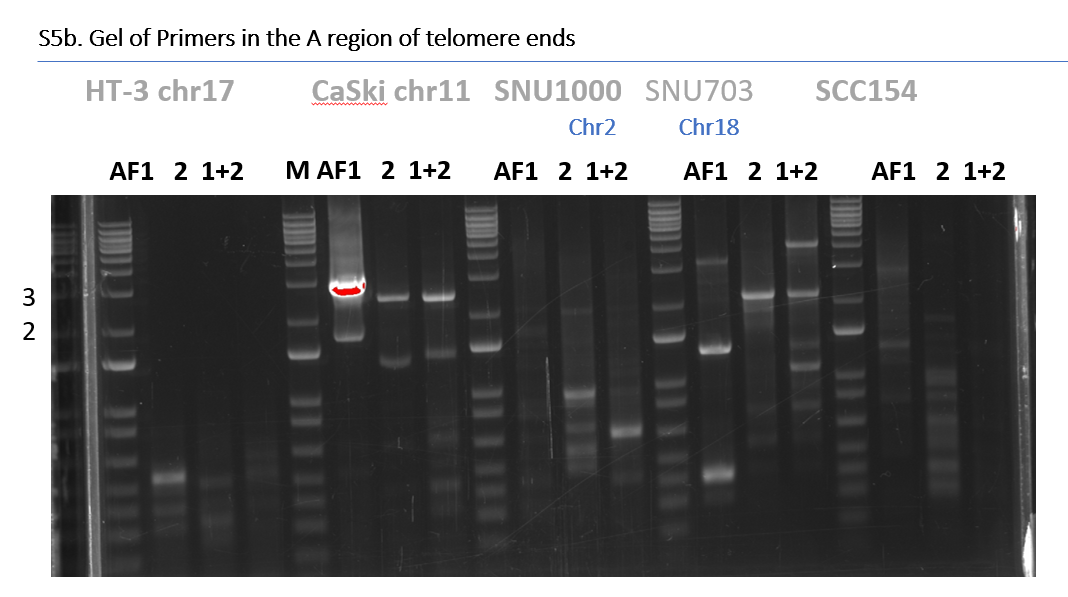

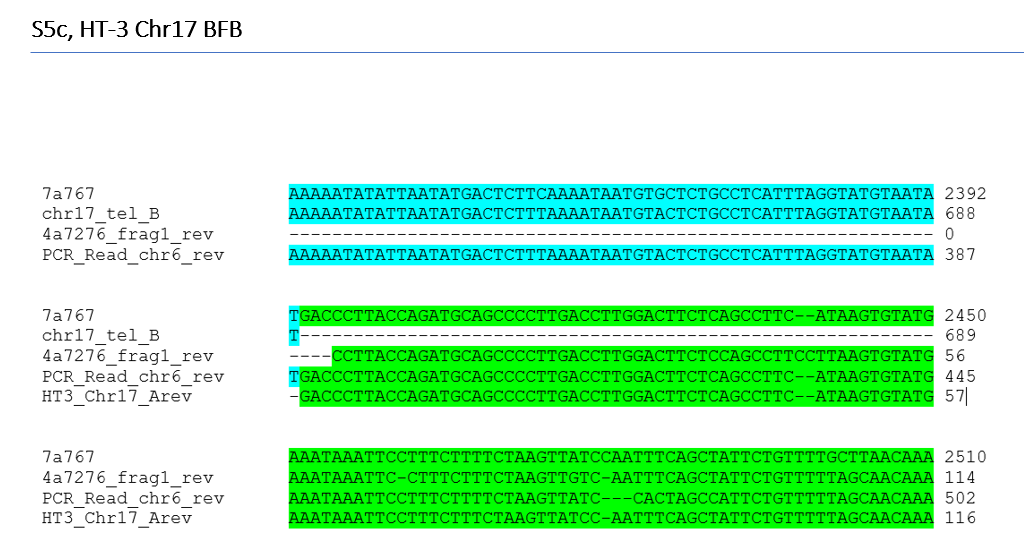

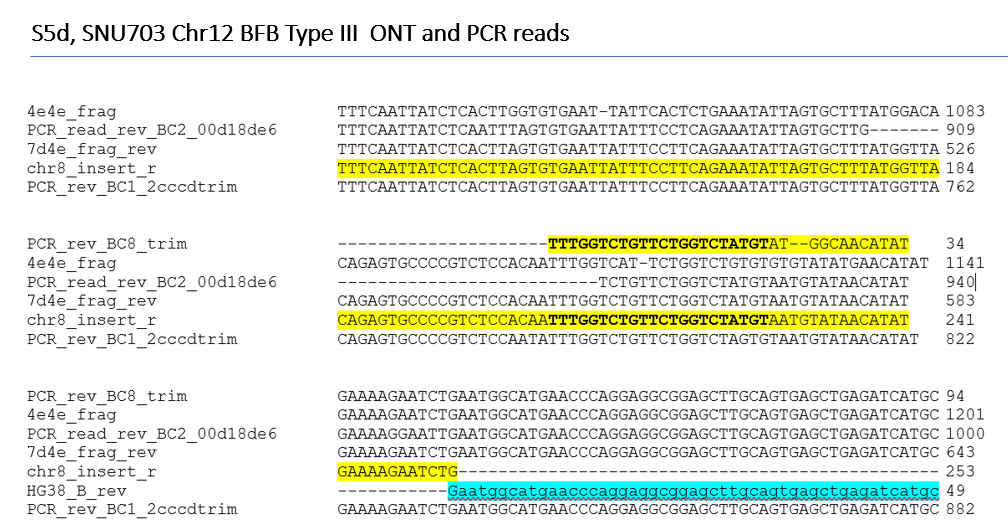

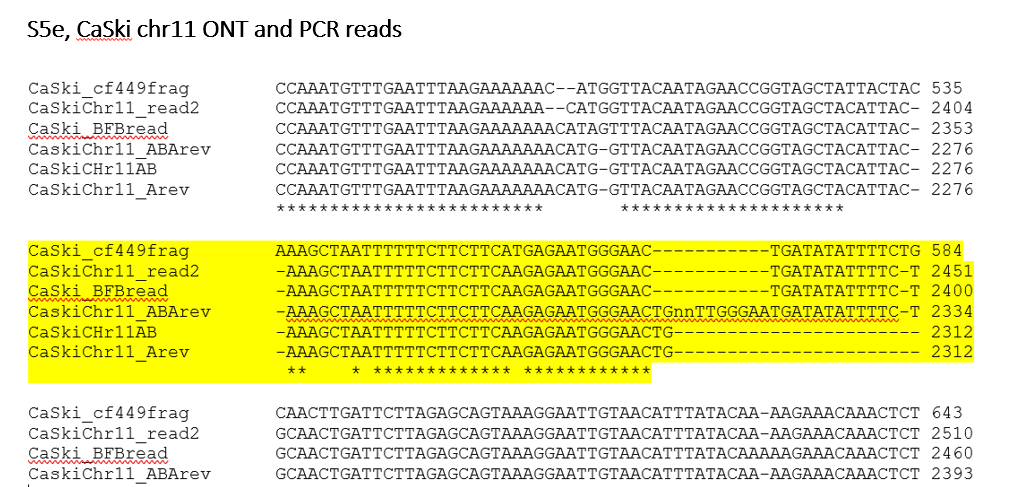

**Figure S5. Strategy for PCR validation of BFB inversions. A**, Using the CaSki chr 11 BFB as an example, primers were designed to single-copy sequencing in segments A and B. Primers to segment A should amplify in a single-primer reaction if there is an inversion and primers in B should amplify bands with primers A. **B.** Examples of a gel displaying the amplification products employing primer AF1 alone, AF2 alone (2) or AF1 and 2 (1+2). The position of the 2 and 3 kb markers are shown on the left. **C,** alignment of the sequence of ONT reads and PCR products for the chr 17 BFB event in HT-3 cells. Segment B is in blue, and segment A in green. **D,** ONT and PCR reads for chr12 in SNU-703 with the chr8 insertion in yellow and segment B in blue. The position of the primer is in bold. E, Alignment of ONT and PCR reads to from CaSki chr11. Highlighted is the junction region.

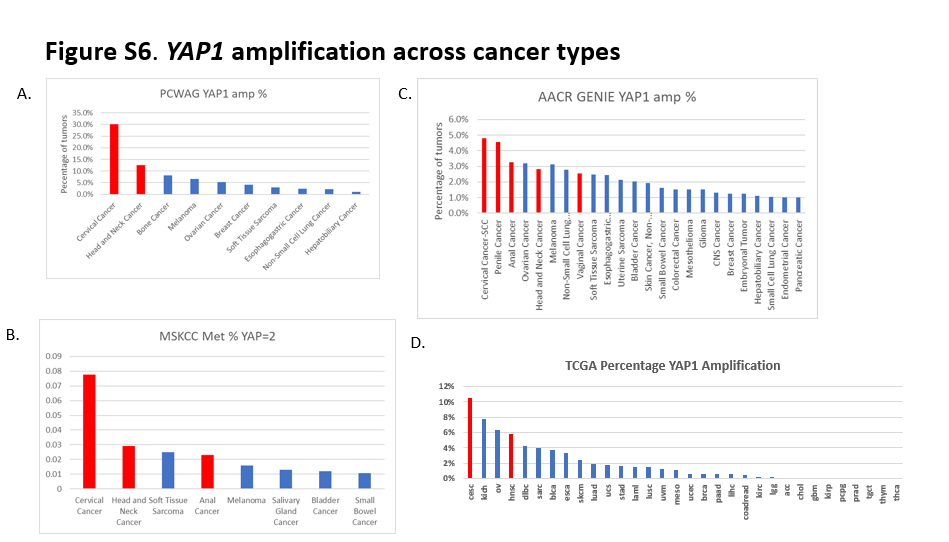

**Figure S6. Frequency of YAP1 amplification across cancer types.** The frequency of *YAP1* gene amplification is shown for HPV-associated cancers (red) and others (blue). **A,** the Pan-cancer analysis of whole genomes (PCAWG). **B,** The Memorial Sloan Kettering metastasis study (MSKCC met), **C,** the AACR GENIE cohort. **D,** TCGA Pan-Cancer cohort.
